# Association of COVID-19 vaccines ChAdOx1 and BNT162b2 with major venous, arterial, or thrombocytopenic events: whole population cohort study in 46 million adults in England

**DOI:** 10.1101/2021.08.18.21262222

**Authors:** CVD-COVID-UK consortium, Writing committee, William N Whiteley, Samantha Ip, Jennifer A Cooper, Thomas Bolton, Spencer Keene, Venexia Walker, Rachel Denholm, Ashley Akbari, Efosa Omigie, Sam Hollings, Emanuele Di Angelantonio, Spiros Denaxas, Angela Wood, Jonathan A C Sterne, Cathie Sudlow

## Abstract

**Background:** Thromboses in unusual locations after the COVID-19 vaccine ChAdOx1-S have been reported. Better understanding of population-level thrombotic risks after COVID-19 vaccination is needed.

**Methods:** We analysed linked electronic health records from adults living in England, from 8^th^ December 2020 to 18^th^ March 2021. We estimated incidence rates and hazard ratios (HRs) for major arterial, venous and thrombocytopenic outcomes 1-28 and >28 days after first vaccination dose for ChAdOx1-S and BNT162b2 vaccines. Analyses were performed separately for ages <70 and ≥70 years, and adjusted for age, sex, comorbidities, and social and demographic factors.

**Results:** Of 46,162,942 adults, 21,193,814 (46%) had their first vaccination during follow-up. Adjusted HRs 1-28 days after ChAdOx1-S, compared with unvaccinated rates, at ages <70 and ≥70 respectively, were 0.97 (95% CI: 0.90-1.05) and 0.58 (0.53–0.63) for venous thromboses, and 0.90 (0.86-0.95) and 0.76 (0.73-0.79) for arterial thromboses. Corresponding HRs for BNT162b2 were 0.81 (0.74–0.88) and 0.57 (0.53–0.62) for venous thromboses, and 0.94 (0.90-0.99) and 0.72 (0.70-0.75) for arterial thromboses. HRs for thrombotic events were higher at younger ages for venous thromboses after ChAdOx1-S, and for arterial thromboses after both vaccines.

Rates of intracranial venous thrombosis (ICVT) and thrombocytopenia in adults aged <70 years were higher 1-28 days after ChAdOx1-S (adjusted HRs 2.27, 95% CI:1.33– 3.88 and 1.71, 1.35–2.16 respectively), but not after BNT162b2 (0.59, 0.24–1.45 and 1.00, 0.75–1.34) compared with unvaccinated. The corresponding absolute excess risks of ICVT 1-28 days after ChAdOx1-S were 0.9–3 per million, varying by age and sex.

**Conclusions:** Increases in ICVT and thrombocytopenia after ChAdOx1-S vaccination in adults aged <70 years were small compared with its effect in reducing COVID-19 morbidity and mortality, although more precise estimates for adults <40 years are needed. For people aged ≥70 years, rates of arterial or venous thrombotic, events were generally lower after either vaccine.

## INTRODUCTION

In late February 2021 several groups reported a rare syndrome of thrombosis and thrombocytopenia after vaccination against severe acute respiratory syndrome coronavirus-2 disease 2019 (COVID-19) with ChAdOx1-S (developed by Oxford-AstraZeneca).^1–3^ Thromboses were found in unusual sites such as the cerebral venous sinuses, or mesenteric or portal veins. This syndrome, named ‘vaccine-induced immune thrombotic thrombocytopenia’ (VITT), is probably due to an autoimmune response to platelet factor 4 (PF4) in the absence of exposure to heparin.^1^ Whether the risk of common thrombotic illnesses is also increased post-vaccination is uncertain, although there have been reports of post-vaccination ischaemic stroke and intracerebral haemorrhage.^1,3,4^

Estimates of vaccine-associated excess risk based on case reports and routine reporting may be biased if diagnostic thresholds vary between vaccinated and unvaccinated individuals or there is less than population-wide coverage. In England, the National Health Service (NHS) provides almost all healthcare; therefore, linked health care records provide comprehensive, population-wide data on outcomes before and after vaccination.^5^ Vaccination in England began on December 8^th^ 2020: the first groups vaccinated were care home residents, then those aged ≥80 years and frontline health and social care workers. Those clinically extremely vulnerable were invited alongside people aged ≥70 years, and those with other co-morbidities aged <65 alongside people aged >65 years. Consequently, individuals vaccinated earlier are expected to have higher rates of venous and arterial events.

We conducted a cohort study using routinely collected, linked health data from multiple sources, covering almost the entire English adult population (>46 million). Associations of ChAdOx1-S and BNT162b2 (developed by Pfizer–BioNTech) with major venous and arterial events and thrombocytopenic haematological outcomes were estimated from the start of the vaccination program to mid-March 2021, before VITT surveillance was widespread.

## METHODS

### Population

The study population was adults (aged ≥18 years), alive and registered with an English NHS general practice on December 8th 2020. The data resource^5^ includes primary care data (General practice extraction service (GPES) data for Pandemic Planning and Research, GDPPR) from 98% of general practices linked at individual-level to nationwide secondary care data including all NHS hospital admissions (Hospital Episode Statistics (HES) and Secondary Uses Service (SUS) data from 1997 onwards), COVID-19 laboratory testing data, COVID-19 vaccination data (NHS England immunisation management system), national community drug dispensing data (NHS BSA Dispensed Medicines from 2018) and death registrations. We accessed and analysed pseudonymised data within NHS Digital’s secure, privacy protecting Trusted Research Environment.^6^

### Covariates

Covariates were defined from primary care, hospital admissions, community drug dispensing and COVID-19 laboratory testing data, using phenotyping algorithms verified by specialist physicians (see study protocol). Phenotypes for co-morbidities, risk factors and other covariates used Systematized Nomenclature of Medicine Clinical Terms (SNOMED-CT) concepts for primary care data, and ICD-10 codes for hospital admission data. Sex, age, region, deprivation and smoking status were defined as the latest recorded in primary care records before 8^th^ December 2020, and the most recently recorded ethnicity in either primary care or hospital admissions records was used. History of diabetes, depression, obesity, cancer, chronic obstructive pulmonary disease, chronic kidney disease, stroke, MI, DVT or PE, cancer, thrombophilia, liver disease or dementia were defined as any record in primary care and/or hospital admission data before December 8th 2020. The number of unique diseases in SNOMED-CT for the year before December 8^th^ 2020 were derived from primary care records and major surgery in the previous year from hospital admission records (using the Office of Population Censuses and Surveys classification of surgical procedures).

A history of coronavirus (SARS-CoV2) infection was defined as either a positive COVID-19 antigen test in national laboratory data covering swab tests performed in the general population and hospitals or a confirmed COVID-19 diagnosis in primary care or hospital admission records. Prior medication was defined using community dispensing data on all prescriptions dispensed by community pharmacists, appliance contractors, and doctors in England. British National Formulary (BNF) codes were used to define the total number of types of medication prescribed and dispensed in the following groups: antiplatelets, antihypertensives, lipid lowering agents, oral anticoagulants, combined oral contraceptives, and hormone replacement therapy.

### Outcomes

Outcomes were derived from primary care data, hospital admission data (SUS dataset) and the national death registry (see supplementary material).^7^ Specialist clinician-verified SNOMED-CT and ICD-10 rule-based phenotyping algorithms were used to define fatal or non-fatal (i) arterial thrombotic events: myocardial infarction (MI), ischaemic stroke (ischaemic or unclassified stroke, spinal stroke or retinal infarction), other non-stroke non-MI arterial thrombo-embolism; (ii) venous thrombo-embolic events: pulmonary embolism (PE), lower limb deep venous thrombosis (DVT), intracranial venous thrombosis (ICVT), portal vein thrombosis, and venous thrombosis at other sites; (iii) thrombocytopenic haematological events: any thrombocytopenia (idiopathic, primary, secondary or unspecified), disseminated intravascular coagulation (DIC), and thrombotic thrombocytopenic purpura (TTP); (iv) death from any cause; (v) other vascular outcomes: haemorrhagic stroke (intracerebral or subarachnoid), and mesenteric thrombus for which available codes did not distinguish between arterial or venous causes; and (vi) lower limb fracture as a control condition unlikely to be affected by vaccination. The event date was the earliest of: start of hospital episode, death, or recorded date of primary care event or consultation. Events within the death registry were identified based on underlying cause of death and in hospital admission data based on the primary cause of the care episode.

### Statistical analyses

Data were analysed according to a pre-specified plan, published on GitHub on April 27^th^ 2021.^8^ Follow-up was from December 8^th^ 2020 to March 18^th^ 2021, the date the European Medicine Agency (EMA) Pharmacovigilance Risk Assessment Committee (PRAC) discussed the first reported complications of vaccination, after which diagnostic effort is expected to be concentrated in people receiving ChAdOx1-S.^9^ Any cell numbers <5 and any potentially disclosive numbers <10 are not reported exactly but as <5 and <10 respectively.

Follow up time for each person was split into periods before, and 1-28 and >28 days after, first vaccination. Censoring was at the earliest of the outcome, death, March 18^th^ 2021 and, in analyses of specific vaccines, receipt of the other vaccine. Incidence rates per 100,000 people per year were estimated for each outcome, before and 1-28 and >28 days after vaccination.

Cox models with time zero December 8^th^ 2020 were fitted, ensuring that all analyses accounted for changes in rates of outcome events with calendar time. Hazard ratios comparing the periods 1-28 and >28 days after vaccination with unvaccinated or pre-vaccination person-time (reference) were estimated. Cox models were fitted separately by age group (<70 and ≥70 years), both overall and separately for males and females. All Cox models were stratified by geographic region. For rare outcomes, sex-specific hazard ratios were estimated from vaccine-sex interaction terms. For computational efficiency, each model included data from all people with, and a 10% random sample of people without, the outcome of interest; analyses incorporated inverse probability weights to account for this sampling. Confidence intervals were derived using robust standard errors.

Region-; age-sex-region-; and fully-adjusted hazard ratios (HR) for associations of ChAdOx1-S and BNT162b2 with outcome events were estimated. The following covariates were included in all fully-adjusted models: (a) sex, age and age^2^, ethnicity, postcode-derived deprivation level; (b) risk factors for venous thromboses (anticoagulant medication, combined oral contraceptive medication, hormone replacement therapy medication, history of PE or DVT, and history of coronavirus infection); (c) risk factors for arterial thromboses (diabetes, hypertension, smoking, antiplatelet medication, blood pressure lowering medication, lipid lowering medication, anticoagulant medication, history of stroke, and history of MI); (d) further covariates selected using a backwards stepwise procedure with p value threshold 0.2, from models using MI as outcome (separately in the four groups defined by age and sex). The covariates had few missing values (apart from ethnicity, for which the 5.9% ‘missing’ were included as a separate category); hence all analyses used “complete-cases”.

Sensitivity analyses examined outcome events recorded as the primary or secondary reason for admission or death in hospital admissions or death records; fatal outcome events (those followed by death from any cause within 28-days); and all venous and all arterial thrombotic outcome events associated with thrombocytopenia. Effect modification by age, sex, ethnicity, medication, diabetes, deprivation and medical history was examined for all venous and all arterial thromboses.

Absolute excess risks of ICVT were estimated by applying estimated hazard ratios to incidence rates of first fatal or non-fatal events in decades of age and sex in 2019, derived from linked hospital and death records.

The study was approved by the Newcastle & North Tyneside 2 Research Ethics Committee (20/NE/0161), the NHS Digital Data Access Request Service (DARS-NIC-381078-Y9C5K) and the British Heart Foundation Data Science Centre CVD-COVID-UK Approvals and Oversight Board.

Data manipulation and analyses used SQL and Python in Databricks and RStudio (Professional) Version 1.3.1093.1 driven by R Version 4.0.3. All code and phenotypes are available at github.com/BHFDSC/CCU002_02.

## RESULTS

On December 8^th^ 2020, 46,162,942 eligible adults were registered with an English general practice. By March 18^th^ 2021, 21,193,814 (46%) had received their first vaccination (8,712,477 BNT162b2; 12,481,337 ChAdOx1-S) (supplementary figure 1). Post-vaccination person-years of follow up for ages <40, 40-69 and ≥70 years respectively were, for ChAdOx1-S: 91,206, 417,160 and 401,343 person years; for BNT162b2: 159,856, 427,874, and 583, 890 person years.

The risks per 100,000 persons from December 8^th^ 2020 to March 18^th^ 2021 were: any venous thrombosis 45.3, any arterial thrombosis 189; thrombocytopenia 4.2 (Table 1). Risks of venous and arterial thromboses were higher in people with co-morbidities and with increasing age and deprivation, and varied substantially by ethnicity. Absolute and relative increases in risk with increasing age and deprivation were greater for arterial than for venous thromboses. The risk of venous thromboses was higher in people with prior DVT or PE, thrombophilia, or oral anticoagulant medication, while the risk of arterial thromboses was higher in people with prior stroke, MI or antiplatelet medication. The risk of thrombocytopenia increased with age and co-morbidities but varied less markedly with ethnicity and deprivation than the risk of venous or arterial thromboses.

**Table 1.** Numbers of patients analysed and (in parentheses) risk per 100,000 people of venous and arterial events and thrombocytopenia (TCP), overall and according to age.

|  |  | Overall |  |  |  | Age, <70 |  |  |  | Age, ≥70 |  |  |  |
| --- | --- | --- | --- | --- | --- | --- | --- | --- | --- | --- | --- | --- | --- |
|  |  | All | Venous | Arterial | TCP | All | Venous | Arterial | TCP | All | Venous | Arterial | TCP |
| N |  | 46,162,942 | 20,903 (45.3) | 87,251 (189) | 1,926 (4.2) | 38,744,171 | 11,835 (30.5) | 38,283 (98.8) | 1,167 (3.0) | 7,418,771 | 9,068 (122) | 48,968 (660) | 759 (10.2) |
| Sex | Male | 22,765,779 | 10,358 (45.5) | 51,851 (228) | 986 (4.3) | 19,425,454 | 6,406 (33.0) | 26,549 (137) | 555 (2.9) | 3,340,325 | 3,952 (118) | 25,302 (757) | 431 (12.9) |
|  | Female | 23,397,163 | 10,545 (45.1) | 35,400 (151) | 940 (4.0) | 19,318,717 | 5,429 (28.1) | 11,734 (60.7) | 612 (3.2) | 4,078,446 | 5,116 (125) | 23,666 (580) | 328 (8.0) |
| Age | 18 - 29 | 8,821,973 | 781 (8.9) | 400 (4.5) | 143 (1.6) | 8,821,973 | 781 (8.9) | 400 (4.5) | 143 (1.6) |  |  |  |  |
|  | 30 - 49 | 16,131,025 | 3,544 (22.0) | 6,424 (39.8) | 338 (2.1) | 16,131,025 | 3,544 (22.0) | 6,424 (39.8) | 338 (2.1) |  |  |  |  |
|  | 50 - 69 | 13,791,173 | 7,510 (54.5) | 31,459 (228) | 686 (5.0) | 13,791,173 | 7,510 (54.5) | 31,459 (228) | 686 (5.0) |  |  |  |  |
|  | 70 - 79 | 4,674,973 | 4,820 (103) | 22,249 (476) | 418 (8.9) |  |  |  |  | 4,674,973 | 4,820 (103) | 22,249 (476) | 418 (8.9) |
|  | 80+ | 2,743,798 | 4,248 (155) | 26,719 (974) | 341 (12.4) |  |  |  |  | 2,743,798 | 4,248 (155) | 26,719 (974) | 341 (12.4) |
| Ethnicity | Asian or Asian British | 3,718,442 | 599 (16.1) | 5,325 (143) | 152 (4.1) | 3,467,223 | 475 (13.7) | 3,378 (97.4) | 125 (3.6) | 251,219 | 124 (49.4) | 1,947 (775) | 27 (10.7) |
|  | Black or Black British | 1,634,470 | 654 (40.0) | 2,086 (128) | 58 (3.5) | 1,525,603 | 513 (33.6) | 1,384 (90.7) | 45 (2.9) | 108,867 | 141 (130) | 702 (645) | 13 (11.9) |
|  | Mixed | 712,534 | 182 (25.5) | 642 (90.1) | 22 (3.1) | 682,037 | 153 (22.4) | 426 (62.5) | 19 (2.8) | 30,497 | 29 (95.1) | 216 (708) | <5 (9.8) |
|  | Other ethnic groups | 1,497,133 | 217 (14.5) | 1,072 (71.6) | 39 (2.6) | 1,427,187 | 162 (11.4) | 678 (47.5) | 33 (2.3) | 69,946 | 55 (78.6) | 394 (563) | <10 (8.6) |
|  | White | 36,446,855 | 18,995 (52.1) | 77,118 (212) | 1,643 (4.5) | 29,619,868 | 10,362 (35.0) | 31,821 (107) | 939 (3.2) | 6,826,987 | 8,633 (126) | 45,297 (663) | 704 (10.3) |
|  | Unknown or missing | 2,153,508 | 256 (11.9) | 1,008 (46.8) | 12 (0.6) | 2,022,253 | 170 (8.4) | 596 (29.5) | 6 (0.3) | 131,255 | 86 (65.5) | 412 (314) | <10 (4.6) |
| Deprivation<sup>1</sup> | 1 - 2 | 9,118,746 | 4,565 (50.1) | 18,830 (206) | 365 (4.0) | 8,069,275 | 3,064 (38.0) | 10,493 (130) | 272 (3.4) | 1,049,471 | 1,501 (143) | 8,337 (794) | 93 (8.9) |
|  | 3 - 4 | 9,567,633 | 4,105 (42.9) | 17,436 (182) | 370 (3.9) | 8,273,028 | 2,454 (29.7) | 8,370 (101) | 234 (2.8) | 1,294,605 | 1,651 (128) | 9,066 (700) | 136 (10.5) |
|  | 5 - 6 | 9,355,549 | 4,091 (43.7) | 17,642 (189) | 392 (4.2) | 7,784,840 | 2,235 (28.7) | 7,263 (93.3) | 221 (2.8) | 1,570,709 | 1,856 (118) | 10,379 (661) | 171 (10.9) |
|  | 7 - 8 | 9,076,313 | 4,146 (45.7) | 16,939 (187) | 381 (4.2) | 7,362,375 | 2,128 (28.9) | 6,334 (86.0) | 221 (3.0) | 1,713,938 | 2,018 (118) | 10,605 (619) | 160 (9.3) |
|  | 9 - 10 | 8,772,902 | 3,926 (44.8) | 16,024 (183) | 406 (4.6) | 7,000,538 | 1,908 (27.3) | 5,602 (80.0) | 210 (3.0) | 1,772,364 | 2,018 (114) | 10,422 (588) | 196 (11.1) |
| Smoking status | Current | 7,814,045 | 3,079 (39.4) | 16,115 (206) | 260 (3.3) | 7,287,225 | 2,492 (34.2) | 11,811 (162) | 219 (3.0) | 526,820 | 587 (111) | 4,304 (817) | 41 (7.8) |
|  | Former | 10,623,072 | 6,930 (65.2) | 31,768 (299) | 581 (5.5) | 7,826,062 | 3,233 (41.3) | 11,703 (150) | 277 (3.5) | 2,797,010 | 3,697 (132) | 20,065 (717) | 304 (10.9) |
|  | Never | 25,722,494 | 10,667 (41.5) | 38,701 (150) | 1,053 (4.1) | 21,674,974 | 5,929 (27.4) | 14,346 (66.2) | 643 (3.0) | 4,047,520 | 4,738 (117) | 24,355 (602) | 410 (10.1) |
| Medical history | Stroke | 727,218 | 1,020 (140) | 21,489 (2,955) | 104 (14.3) | 286,650 | 323 (113) | 8,147 (2,842) | 40 (14.0) | 440,568 | 697 (158) | 13,342 (3,028) | 64 (14.5) |
|  | Myocardial infarction | 1,189,182 | 1,436 (121) | 24,212 (2,036) | 158 (13.3) | 485,506 | 436 (89.8) | 10,486 (2,160) | 46 (9.5) | 703,676 | 1,000 (142) | 13,726 (1,951) | 112 (15.9) |
|  | DVT or PE | 603,351 | 3,263 (541) | 4,050 (671) | 142 (23.5) | 341,788 | 1,919 (561) | 1,377 (403) | 76 (22.2) | 261,563 | 1,344 (514) | 2,673 (1,022) | 66 (25.2) |
|  | Thrombophilia | 44,593 | 146 (327) | 202 (453) | 23 (51.6) | 38,136 | 126 (330) | 144 (378) | 21 (55.1) | 6,457 | 20 (310) | 58 (898) | <5 (31.0) |
|  | Coronavirus infection<sup>2</sup> | 1,284,984 | 1,131 (88.0) | 4,167 (324) | 101 (7.9) | 1,124,135 | 576 (51.2) | 1,642 (146) | 63 (5.6) | 160,849 | 555 (345) | 2,525 (1,570) | 38 (23.6) |
|  | Diabetes | 4,069,412 | 3,654 (89.8) | 25,595 (629) | 423 (10.4) | 2,460,649 | 1,509 (61.3) | 9,949 (404) | 205 (8.3) | 1,608,763 | 2,145 (133) | 15,646 (973) | 218 (13.6) |
|  | Depression | 9,516,020 | 6,272 (65.9) | 24,146 (254) | 500 (5.3) | 8,040,590 | 4,077 (50.7) | 12,859 (160) | 348 (4.3) | 1,475,430 | 2,195 (149) | 11,287 (765) | 152 (10.3) |
|  | Obesity | 10,918,274 | 8,863 (81.2) | 31,194 (286) | 679 (6.2) | 8,757,004 | 5,430 (62.0) | 15,848 (181) | 411 (4.7) | 2,161,270 | 3,433 (159) | 15,346 (710) | 268 (12.4) |
|  | Cancer | 6,609,691 | 5,674 (85.8) | 17,827 (270) | 786 (11.9) | 4,854,822 | 2,583 (53.2) | 4,462 (91.9) | 412 (8.5) | 1,754,869 | 3,091 (176) | 13,365 (762) | 374 (21.3) |
|  | COPD | 1,582,366 | 2,497 (158) | 12,323 (779) | 182 (11.5) | 714,880 | 877 (123) | 3,839 (537) | 76 (10.6) | 867,486 | 1,620 (187) | 8,484 (978) | 106 (12.2) |
|  | Liver disease | 230,243 | 284 (123) | 1,069 (464) | 97 (42.1) | 173,423 | 208 (120) | 547 (315) | 75 (43.2) | 56,820 | 76 (134) | 522 (919) | 22 (38.7) |
|  | CKD | 2,934,107 | 4,467 (152) | 25,816 (880) | 526 (17.9) | 978,886 | 1,179 (120) | 5,164 (528) | 200 (20.4) | 1,955,221 | 3,288 (168) | 20,652 (1,056) | 326 (16.7) |
|  | Major surgery<sup>3</sup> | 3,995,870 | 5,407 (135) | 24,164 (605) | 763 (19.1) | 2,773,174 | 2,735 (98.6) | 9,898 (357) | 440 (15.9) | 1,222,696 | 2,672 (219) | 14,266 (1,167) | 323 (26.4) |
|  | Dementia | 524,293 | 1,127 (215) | 6,073 (1,158) | 45 (8.6) | 48,821 | 81 (166) | 415 (850) | 10 (20.5) | 475,472 | 1,046 (220) | 5,658 (1,190) | 35 (7.4) |
| Medications | Antiplatelet | 2,510,382 | 2,814 (112) | 36,415 (1,451) | 218 (8.7) | 1,000,010 | 747 (74.7) | 15,556 (1,556) | 65 (6.5) | 1,510,372 | 2,067 (137) | 20,859 (1,381) | 153 (10.1) |
|  | BP lowering | 8,589,860 | 7,969 (92.8) | 55,462 (646) | 777 (9.0) | 4,286,561 | 2,779 (64.8) | 20,579 (480) | 305 (7.1) | 4,303,299 | 5,190 (121) | 34,883 (811) | 472 (11.0) |
|  | Lipid lowering | 6,808,408 | 5,796 (85.1) | 47,859 (703) | 564 (8.3) | 3,212,600 | 1,878 (58.5) | 19,543 (608) | 194 (6.0) | 3,595,808 | 3,918 (109) | 28,316 (787) | 370 (10.3) |
|  | Anticoagulant | 1,338,585 | 2,278 (170) | 12,758 (953) | 205 (15.3) | 381,588 | 1,144 (300) | 2,773 (727) | 57 (14.9) | 956,997 | 1,134 (118) | 9,985 (1043) | 148 (15.5) |
|  | Oral contraceptive | 622,529 | 138 (22.2) | 42 (6.7) | 8 (1.3) | 622,529 | 138 (22.2) | 42 (6.7) | 8 (1.3) |  |  |  |  |
|  | HRT | 540,722 | 213 (39.4) | 516 (95.4) | 16 (3.0) | 501,957 | 185 (36.9) | 368 (73.3) | 14 (2.8) | 38,765 | 28 (72.2) | 148 (382) | <5 (5.2) |
| Number of diagnoses | 0 | 36,455,307 | 12,200 (33.5) | 38,617 (106) | 980 (2.7) | 31,819,272 | 7,612 (23.9) | 18,316 (57.6) | 643 (2.0) | 4,636,035 | 4,588 (99.0) | 20,301 (438) | 337 (7.3) |
|  | 1 - 5 | 9,573,068 | 8,422 (88.0) | 46,012 (481) | 925 (9.7) | 6,872,479 | 4,137 (60.2) | 19,183 (279) | 515 (7.5) | 2,700,589 | 4,285 (159) | 26,829 (993) | 410 (15.2) |
|  | 6+ | 134,567 | 281 (209) | 2,622 (1948) | 21 (15.6) | 52,420 | 86 (164) | 784 (1496) | 9 (17.2) | 82,147 | 195 (237) | 1,838 (2,237) | 12 (14.6) |
| Number of medications | 0 | 22,970,920 | 3,679 (16.0) | 9,629 (41.9) | 321 (1.4) | 22,122,043 | 3,143 (14.2) | 7,258 (32.8) | 290 (1.3) | 848,877 | 536 (63.1) | 2,371 (279) | 31 (3.7) |
|  | 1 - 5 | 20,875,516 | 12,872 (61.7) | 56,255 (269) | 1,210 (5.8) | 15,618,756 | 7,203 (46.1) | 25,416 (163) | 713 (4.6) | 5,256,760 | 5,669 (108) | 30,839 (587) | 497 (9.5) |
|  | 6+ | 2,316,506 | 4,352 (188) | 21,367 (922) | 395 (17.1) | 1,003,372 | 1,489 (148) | 5,609 (559) | 164 (16.3) | 1,313,134 | 2,863 (218) | 15,758 (1,200) | 231 (17.6) |

<sup>1</sup> Index of Multiple Deprivation deciles where 10 indicates least deprived and 1 indicates most deprived; <sup>2</sup> After 31/12/2019 and prior to 08/12/2020; <sup>3</sup> In the last year.

Incidence rates of all events varied substantially in unvaccinated and post-vaccination (Table 2). In the unvaccinated, the crude incidence rates per 100,000 person years were: all venous thromboses 141 (mostly PE and DVT), ICVT 1.97; portal vein thrombosis 1.15; any arterial thrombosis 549 (with slightly more ischaemic stroke (273) than MI (268)); any thrombocytopenia 13, with very low rates of DIC (0.12) and TTP (0.67). For each vaccine, unadjusted incidence rates of venous and arterial thromboses, haemorrhagic stroke, mesenteric thrombus and thrombocytopenia were higher after than before vaccination in the under 70s, but lower after than before vaccination in the over 70s.

**Table 2(a).**
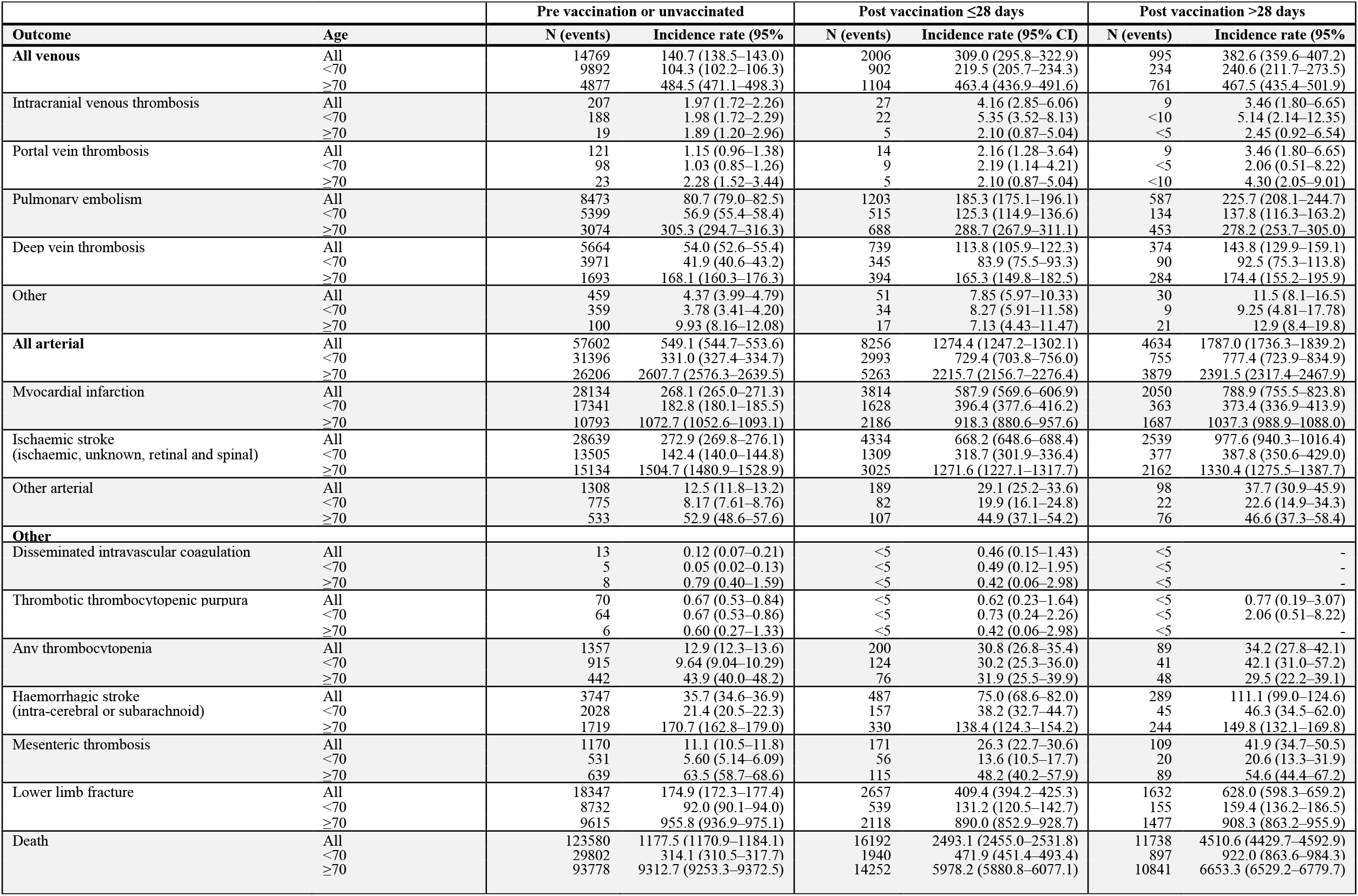
**Numbers and incidence rates pre-, post-first ChAdOx1-S. Incidence rate per 100,000 person years. Disclosure control prevents presentation of n>5**

**Table 2(b).**
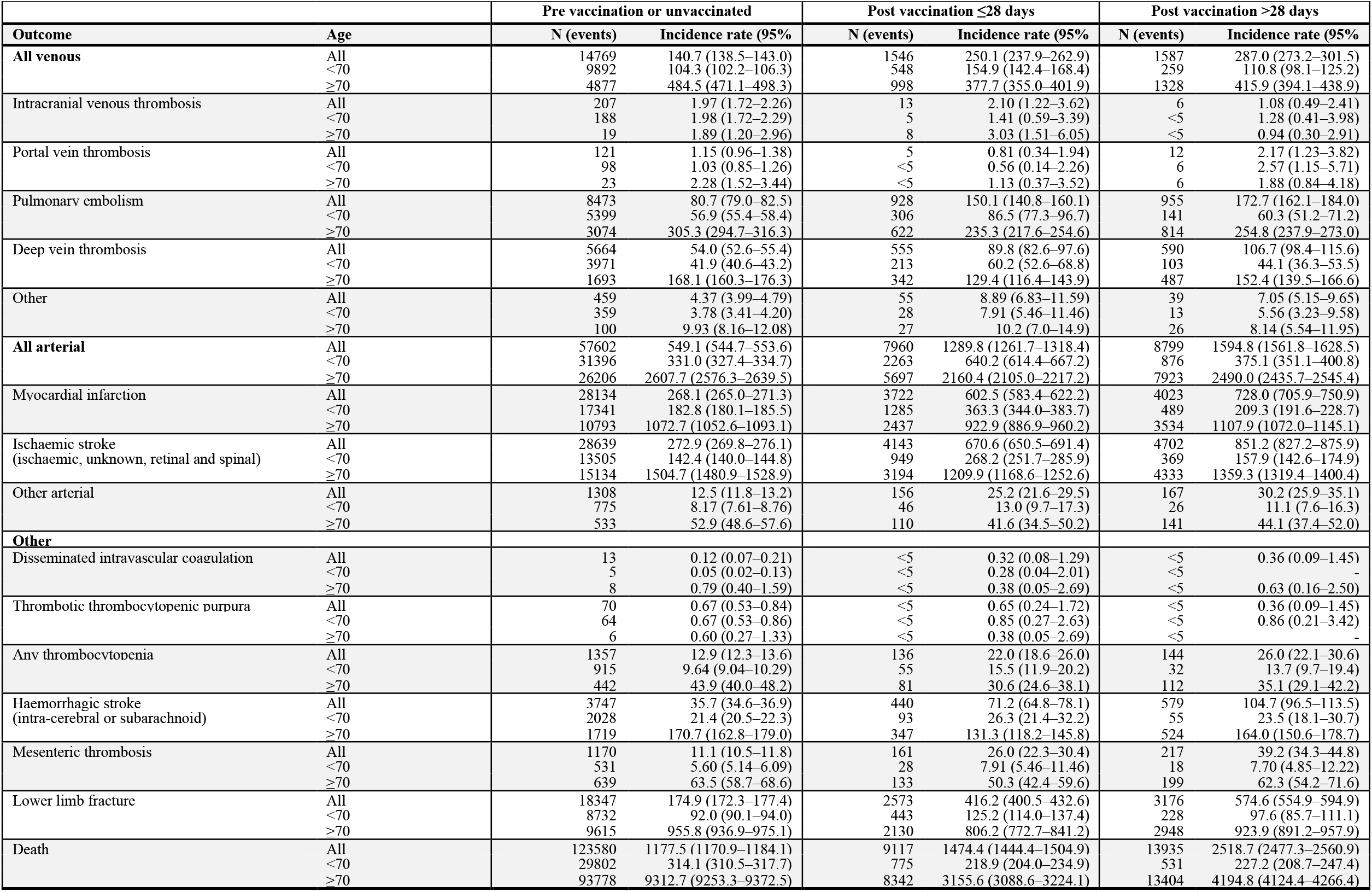
**Numbers and incidence rates pre-, post-first BNT162b2. Incidence rate per 100**,**000 person years. Disclosure control prevents presentation of n>5**

The additional confounding factors selected using backward selection were: for both sexes and age strata, previous diagnosis of cancer, number of unique diseases in the last year, surgery in the last year, obesity, liver disease; additionally for women aged <70, history of depression and total number of types of medication (by BNF chapters); additionally for men aged <70, history of SARS-CoV2 infection and thrombocytopenia; and additionally for men and women aged ≥70, chronic kidney disease and dementia.

Crude hazard ratios were substantially attenuated after adjusting for age, and further attenuated after adjusting for confounding factors. For example, in <70 year-olds the HRs 1-28 days post-versus unvaccinated were attenuated from 2.81 to 2.27 for ICVT; from 4.85 to 1.71 for thrombocytopenia; and from 3.21 to 0.90 for ischaemic stroke (supplementary table 1).

In people aged <70 years, the fully adjusted HR for any venous thrombosis was 0.97 (95% CI:0.90-1.05) 1–28 days after ChAdOx1-S compared with before vaccination, with similar adjusted HRs for PE and DVT. Compared with before vaccination, the hazard of any venous thrombosis was lower 1-28 days after ChAdOx1-S in people aged ≥70 (adjusted HR 0.58, 95%CI:0.53–0.63) and 1–28 days after BNT162b2 (<70: HR 0.81, 0.74–0.88; ≥70 HR: 0.57, 0.53–0.62), with similar adjusted HRs for PE and DVT. (Figure 1, supplementary table 1).

**Figure 1.**
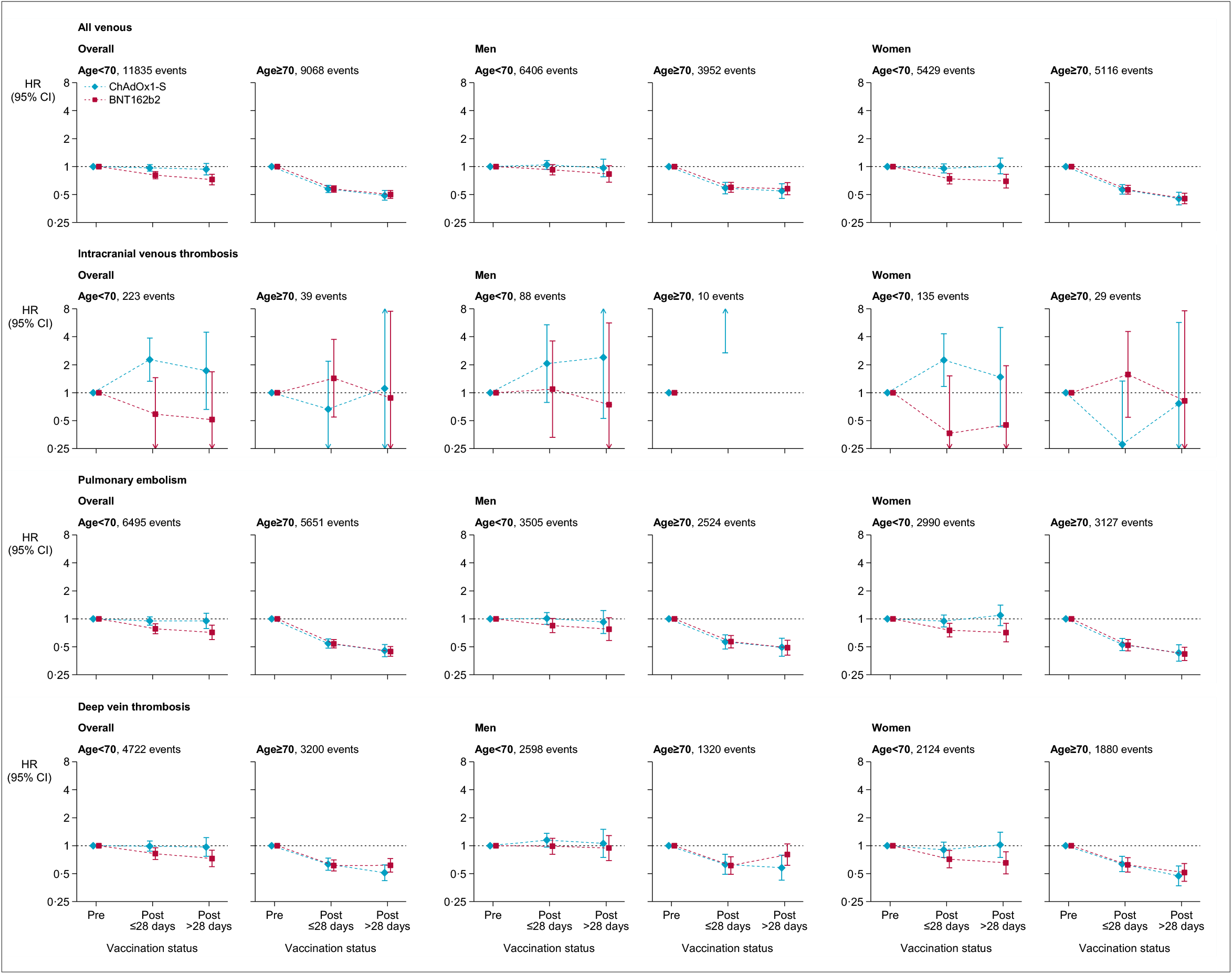
Adjusted Hazard ratios for all venous thromboses, intracranial venous thromboses, pulmonary embolism and deep vein thromboses after ChAdOx1-S or BNT162b2 vaccine

In people aged <70 years, adjusted HRs for ICVT were 2.27 (95%CI: 1.32–3.88) 1-28 days after ChAdOx1-S and 0.51 (0.16–1.68) 1-28 days after BNT162b2. HRs were similar in men and women. In a post-hoc analyses, in people aged <40 years, the HRs 1-28 days after vaccination were 3.73 (1.58–8.83) and 1.07 (0.33–3.48) for ChAdOx1-S and BNT162b2 respectively. The same estimates for people 40-69 years were 1.93 (1.19–2.13) and 0.74 (0.29–1.87). In people aged ≥70 years, associations of vaccination with ICVT were estimated imprecisely (<10 events for ChAdOx1-S and 8 for BNT162b2 (Figure 1, supplementary table 1).

Adjusted HRs for any arterial thrombosis 1-28 days after ChAdOx1-S were 0.90 (95%CI: 0.86-0.95) and 0.76 (0.73–0.79) in people aged <70 and ≥70 years respectively. Corresponding HRs 1-28 days after BNT162b2 were 0.94 (0.90-0.99) and 0.72 (0.70–0.75). HRs were similar for MI and ischaemic stroke, and >28 days after vaccination (Figure 2, supplementary table 1).

**Figure 2.**
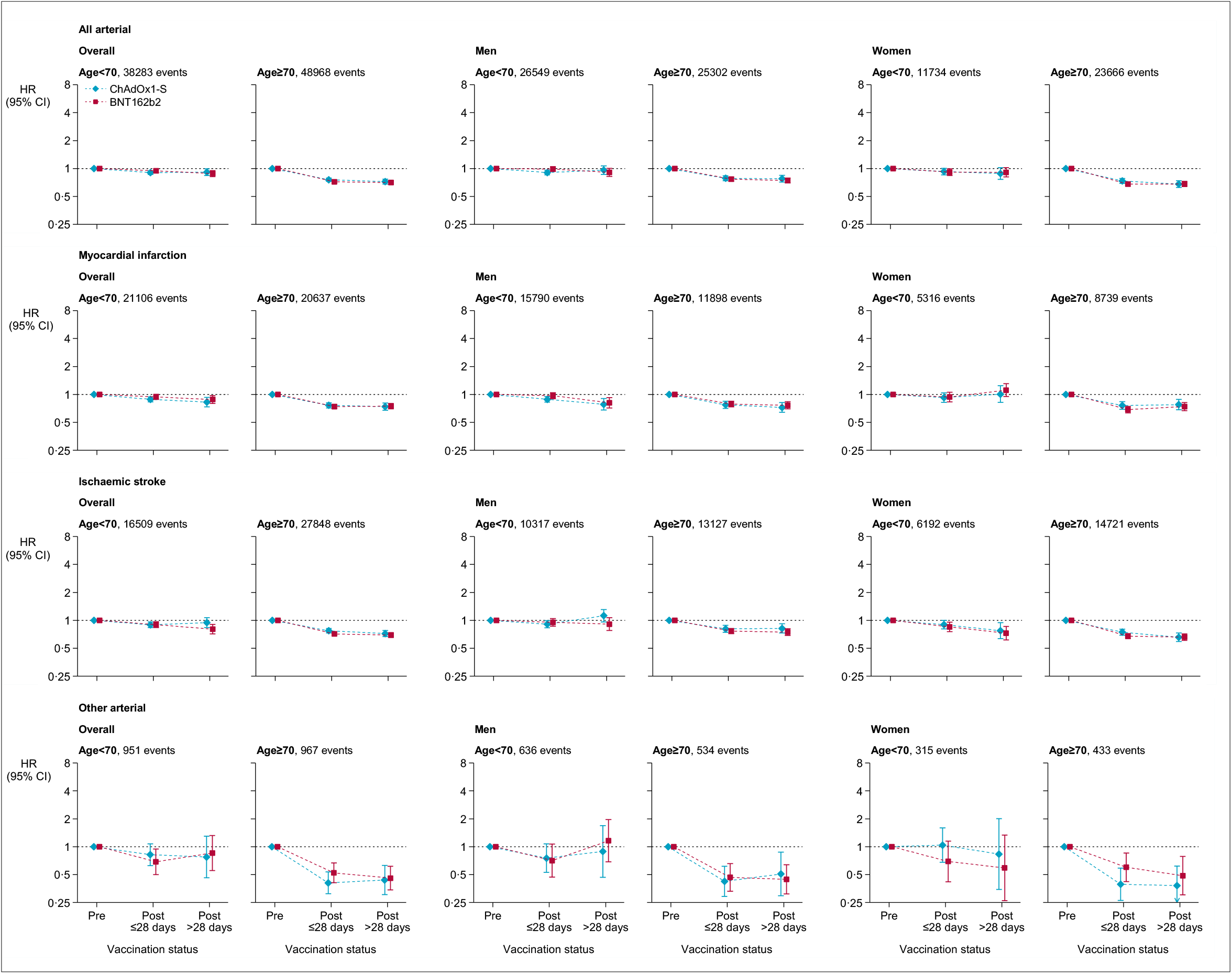
Adjusted hazard ratios for all arterial thromboses, myocardial infarction, ischemic stroke and other arterial thromboses after ChAdOx1-S or BNT162b2 vaccine

In people aged <70 years, adjusted HRs for thrombocytopenia were 1.71 (95%CI:1.35–2.16) 1–28 days and 1.69 (1.16–2.46) >28 days after ChAdOx1-S. Corresponding HRs after BNT162b2 were 1.00 (0.75–1.34) and 0.97 (0.66–1.41). For those aged ≥70 years, adjusted HRs for thrombocytopenia 1-28 days after vaccination were similar for ChAdOx1 (0.79, 95%CI: 0.56–1.10) and BNT162b2 (0.68, 0.51-0.90) (Figure 3, supplementary table 1).

**Figure 3.**
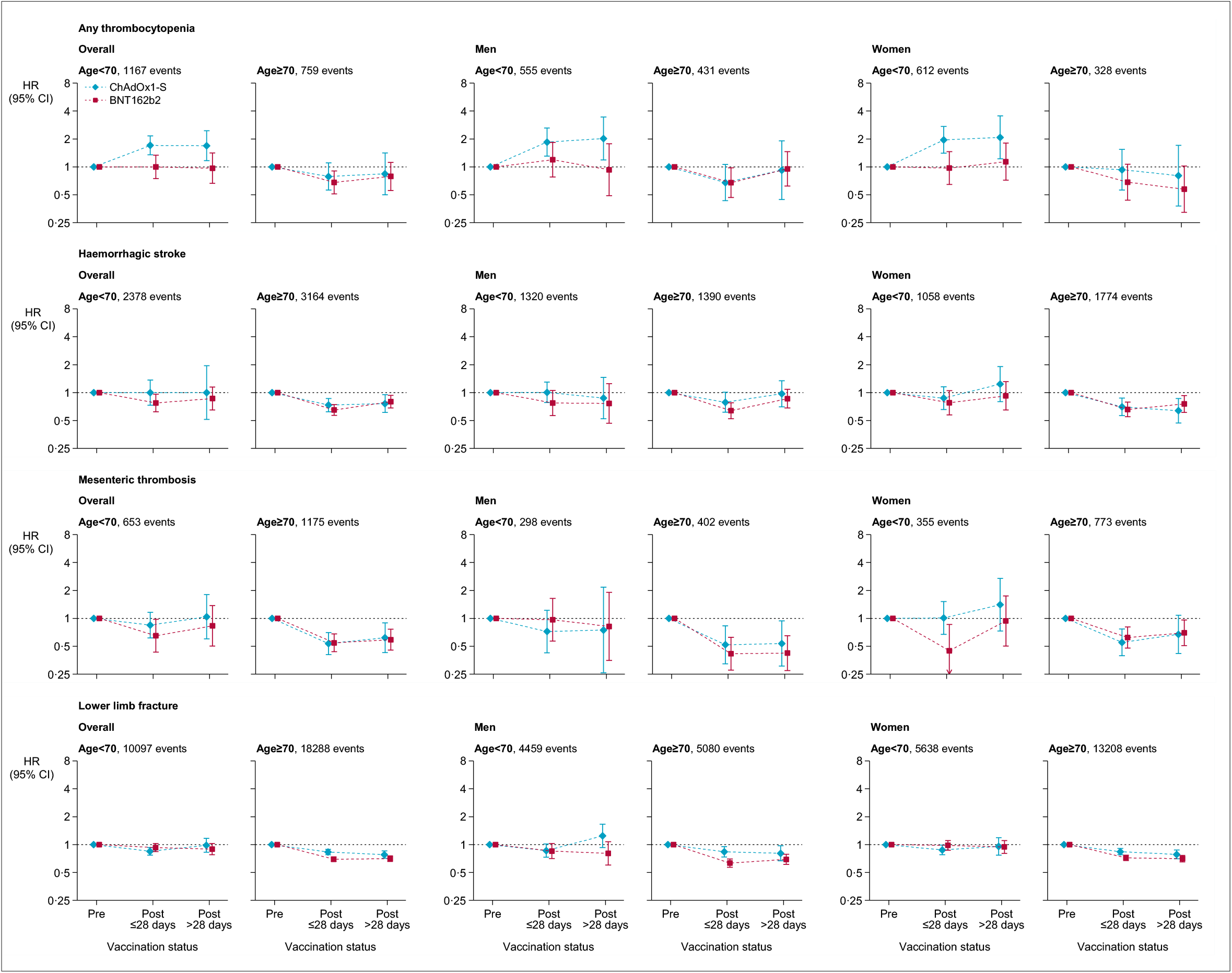
Adjusted hazard ratios for thrombocytopenia hemorrhagic stroke, mesenteric thrombosis, lower limb fracture after ChAdOx1-S or BNT162b2 vaccine

Adjusted HRs for lower limb fracture 1–28 days after ChAdOx1-S were 0.85 (95% CI: 0.77–0.93), and 0.83 (0.78–0.89) in people aged <70 and ≥70 years respectively. Corresponding HRs after BNT162b2 were 0.93 (0.84–1.02) and 0.69 (0.66–0.73). (Figure 3, supplementary table 1). Adjusted HRs for all-cause mortality 1–28 days after ChAdOx1-S were 0.37 (95%CI: 0.35–0.39), and 0.28 (0.28–0.29) in people aged <70 and ≥70 years respectively. Corresponding HRs after BNT162b2 were 0.24 (0.22– 0.26) and 0.19 (0.19–0.20) (supplementary figure 2, supplementary table 1).

### Subgroup analyses

HRs for all venous and for all arterial thrombotic events 1-28 days after vaccination were lower in older age groups for each vaccine (all interaction p values <0.0001) (supplementary table 2). HRs for venous thromboses were >1 1-28 days after compared with before vaccination in people aged <50 years for ChAdOx1-S but not BNT162b2. HRs for arterial thromboses were >1 1-28 days after compared with before vaccination in people aged <50 years for both ChAdOx1-S and BNT162b2. A higher proportion of venous thromboses in those aged <40 years compared with those aged ≥40 years were due to ICVT. A higher proportion of arterial thromboses at age <40 was due to stroke than at ages 40–59 and 60–69 years. Less marked trends for lower HRs in older individuals were observed for HRs >28 days after vaccination. Despite modest differences between the magnitude of HRs in subgroups defined by sex, ethnicity, prior COVID, COCP, diabetes, deprivation or history of MI, DVT or PE, anticoagulation or antiplatelet medication, HRs in all of these subgroups were consistent with lower rates of arterial or venous thromboses after vaccination with ChAdOx1-S or BNT162b2.

### Sensitivity analyses

When using a less restrictive outcome definition (outcomes recorded as primary or secondary reason for admission), estimated HRs were consistent with those from analyses of outcomes in the primary position (supplementary figure 3). Similarly, estimated HRs for fatal outcomes, were similar to HRs for outcomes in the primary position (supplementary figure 4). There were very few people for whom outcomes in the primary position were recorded as well as thrombocytopenia in any position in the same hospital admission or death record (after ChAdOx1-S and BNT162b respectively: ICVT 7 and 0; all arterial thromboses 47 and 58; all venous thromboses 37 and 26).

### Intracranial venous thrombosis

Characteristics were similar among patients who developed ICVT while unvaccinated, after ChAdOx1 and after BNT162b. Most were women (66% unvaccinated, 67% ChAdOx1, 79% BNT162b), of white ethnicity (75% unvaccinated, 92% ChAdOx1, 100% BNT162b), and had no prior co-morbidities (71% unvaccinated, 50% ChAdOx1, 50% BNT162b). Patients with post vaccination ICVT were older than those with unvaccinated ICVT. Post-vaccination, no patients had a recorded history of thrombophilia, and fewer than 5% had a recorded dispensed prescription of oral contraceptive or HRT (supplementary Table 3).

Applying the HR for ICVT 1-28 days after ChAdOx1 to the monthly incidence of ICVT in 2019, the excess risk of ICVT in the month after vaccination with ChAdOx1 among those aged 19–30, 31–39, 40–49, 50–59 and 60–69-years was estimated to be, respectively: 0.9, 1.1, 1.5, 1.5 and 1.6 per million vaccinated in men and 3.0, 2.7, 2.0, 1.6 and 1.5 per million vaccinated in women.

## DISCUSSION

In this cohort study, which included almost all adults alive in England at the start of the national COVID-19 vaccination programme, vaccination with ChAdOx1-S, but not BNT162b2, was associated with approximately 2-fold higher rates of ICVT or hospitalisations due to thrombocytopenia in people aged under 70 years, after adjusting for a comprehensive range of demographic characteristics and comorbidities. The corresponding absolute increases in the risk of these events were very small. ChAdOx1-S and BNT162b2 were associated with lower hazards of major vascular events after vaccination in those aged ≥70 years. Post-hoc analyses stratified by age suggest that in people aged <50 years, rates of venous thromboses were higher 1-28 days after vaccination with ChAdOx1-S, and that rates of arterial thrombotic events were higher 1-28 days after vaccination with ChAdOx1-S or BNT162b2, compared with rates in unvaccinated people.

The large population analysed (including almost 10 times as many vaccinated people as any other published study) adds substantial precision to estimated associations of thromboses with vaccination, which is of particular importance for very rare events and associations in subgroups. The extensive data linkages across different healthcare settings enabled adjustment for a wide range of confounding variables. The analysed population is slightly larger than the mid-year estimate of the 2020 English population aged ≥18 (44,456,850) that excluded short-term residents and students, and modelled international migrant numbers.^10^

Smaller Danish and Norwegian electronic health record studies reported a small excess of intracranial haemorrhage (2 per 100,000 1–28 days after ChAdOx1-S), cerebral venous thrombosis (3 per 100,000) and other venous thrombosis (2 per 100,000).^11^ The randomised trials of ChAdOx1-S, reported no venous thrombotic events among 8597 participants who received the vaccine versus two among 8581who received placebo.^12^ A Scottish study found a risk of of 1.13 (0.62–1.63) cases of idiopathic thrombocytopenic purpura per 100,000 after ChAdOx1-S. There was no clear evidence on other outcomes, although the number of post-vaccine events was small.^13^ Reporting of ICVT cases to the UK Medicines and Healthcare products Regulatory Agency (MHRA) identified more cases of ICVT with thrombocytopenia after ChAdOx1-S than the present study (44 up to 31^st^ March 2021).^14^

Healthcare systems planning to use ChAdOx1-S should balance the very small harms against the known benefits of the vaccine. For older populations, who are most vulnerable to COVID-19, we found no evidence of increased risk of any event with ChAdOx1-S. In younger populations, who have a lower morbidity and mortality due to COVID-19, other available vaccines might be prioritise, especially when the risk of COVID-19 is otherwise low.

Our study has several limitations. First, identification of exposures, covariates and outcomes relies on the accuracy of data collected during routine healthcare. Additional data on results of laboratory and radiology investigations would have improved diagnostic coding, particularly for ascertainment of thrombocytopenia. Second, people who were not registered with an NHS GP (for example the homeless, recent immigrants, those using only private healthcare and those not eligible for NHS care) or who opted out of their data being provided to NHS Digital were excluded. Third, follow up ended on March 18^th^ 2021, but a small number of events that occurred before this date may have been excluded because they had yet to be coded or the people affected were still in hospital. Analyses after this date will likely lead to overestimation of associations, because speciality societies recommended further investigations of mild symptoms in vaccinated populations. Fourth, our primary outcome used the primary reason for death or hospital admission, which improves the positive predictive value but may lead to an underestimation of incidence. This is necessary because historical non-incident events are frequently recorded in secondary positions. Analyses of events recorded in any position as fatal within 28 days were consistent with the primary analyses. Fifth, we did not address time-varying confounding, which can occur when factors which vary during follow up, such as admission to hospital, predict both vaccination and outcomes of interest. Sixth, comparison of adjusted and unadjusted associations suggests that studies that do not adjust for a comprehensive range of potential confounders will overestimate the thrombotic effects of vaccination. However, adjusted associations in this study may still be biased by unmeasured confounding by patient characteristics that predict both vaccination and thromboses and that are difficult to ascertain in electronic health records. Examples include general health at the time of vaccination, and people at higher risk of thromboses (e.g. with end-stage diseases).^15^ There were slightly lower post-vaccination rates of the ‘negative control’ outcome lower limb fracture in those aged ≥70 years, which implies that this degree of unmeasured confounding may affect results for thrombotic outcomes in that age group.

Associations of vaccination with thromboses varied with age. This may be because in older people small increases in the risk of major thrombotic events after vaccination with ChAdOx1-S were more than offset by reductions in major thrombotic events (particularly PE) subsequent to COVID-19. By contrast, in younger people any increase in risk associated with ChAdOx1-S vaccine is less likely to be offset by a lower risk of COVID-associated thrombosis, because the chance of severe COVID disease is lower in younger than older individuals.

Further analyses assessing the effects of other vaccines and the effects of second doses of these vaccines on thrombotic, neurological and cardiac complications will be important to inform vaccination programs. Access to data from radiology or laboratory systems (which in the UK will rely on regional rather than national data collection systems) will allow more comprehensive case ascertainment and more granular phenotyping. Such efforts are currently underway across the UK.

## Data Availability

The de-identified data used in this study is available via the CVD-COVID-UK consortium coordinated by BHF Data Science Centre (https://www.hdruk.ac.uk/projects/cvd-covid-uk-project/) for accredited researchers working on approved projects in the NHS Digital trusted research environment. The study protocol, analytic and phenotyping code is available at: https://github.com/BHFDSC/CCU002_02

## FUNDING

The British Heart Foundation Data Science Centre (grant No SP/19/3/34678, awarded to Health Data Research (HDR) UK) funded co-development (with NHS Digital) of the trusted research environment, provision of linked datasets, data access, user software licences, computational usage, and data management and wrangling support. Support was also provided through the Data and Connectivity and Longitudinal Health and Wellbeing National Core Studies, which were established through the UK Government’s Chief Scientific Adviser’s National Core Studies programme to coordinate COVID-19 priority research. Consortium partner organisations funded the time of contributing data analysts, biostatisticians, epidemiologists, and clinicians.

WW is supported by the Chief Scientist’s Office (CAF/01/17). CS, AW and WW are supported by Stroke Association (SA CV 20\100018). WW has given expert testimony to UK courts.

AMW is supported by the BHF-Turing Cardiovascular Data Science Award (BCDSA\100005) and by core funding from UK MRC (MR/L003120/1), BHF (RG/13/13/30194; RG/18/13/33946), and NIHR Cambridge Biomedical Research Centre (BRC-1215-20014). AMW is part of the BigData@Heart Consortium, funded by the Innovative Medicines Initiative-2 Joint Undertaking under grant agreement No 116074. The views expressed are those of the author(s) and not necessarily those of the NIHR or the Department of Health and Social Care.

JAC, JS, and RD are supported by the Health Data Research (HDR) UK South West Better Care Partnership, and the NIHR Bristol Biomedical Research Centre at University Hospitals Bristol, and Weston NHS Foundation Trust and the University of Bristol.

VMW is supported by the Medical Research Council Integrative Epidemiology Unit, which receives its funding from the Medical Research Council and the University of Bristol (MC_UU_00011/4).

AA, SD is supported by Health Data Research UK (grant number: HDR-9006), which receives its funding from the UK Medical Research Council, Engineering and Physical Sciences Research Council, Economic and Social Research Council, Department of Health and Social Care (England), Chief Scientist Office of the Scottish Government Health and Social Care Directorates, Health and Social Care Research and Development Division (Welsh Government), Public Health Agency (Northern Ireland), British Heart Foundation (BHF) and the Wellcome Trust; and Administrative Data Research UK, which is funded by the Economic and Social Research Council (grant number: ES/S007393/1).

This work uses data provided by patients and collected by the NHS as part of their care and support. We would also like to acknowledge all data providers who make anonymised data available for research.

Funders had no role in the study design, collection, analysis, and interpretation of data; in the writing of the report; and in the decision to submit the article for publication

## Conflicts of interest

None

## CONTRIBITIONS

WW chaired the writing committee. WW, CS, JS and AW conceived the idea, and prepared the protocol and drafted the manuscript. SI, JAC, SK, VW, TB, RD and JS wrote the analytic code. VW, JAC, SK and SI prepared analytic code for open access publication. SI, VW and TB prepared the tables and figures. EO and SH prepared data from NHS Digital. SD, SK and WW prepared phenotype code. AW and JS designed and supervised the statistical analyses. CS is the Director of the BHF Data Science Centre and coordinated approvals for and access to data within the NHS Digital TRE. All authors commented on the protocol, analyses, and manuscript, and approved the final version before submission. All authors had full access to the study data.

## TABLES

Table 1 Numbers of patients analysed and (in parentheses) risk per 100,000 of venous and arterial events and thrombocytopenia (TCP) during follow up, overall and according to age.

Table 2 Numbers of events and incidence rates pre and post first vaccination. Incidence rate per 100,000 per annum (a) ChAdOx1-S (b) BNT162b2 vaccine

## SUPPLEMENTARY MATERIAL

### Protocol

**Supplementary table 1** Unadjusted, age-, sex- and region-adjusted, and fully adjusted hazard ratios for (a) ChAdOx1-S (b) BNT162b2 vaccine

**Supplementary table 2** Stratum specific estimates for (a) venous and (b) arterial events.

**Supplementary table 3**. Characteristics of patient who had an ICVT event before and after vaccination. Only percentage are presented, as disclosure control does not allow numbers <5 to be released

**Supplementary figure 1:** Cumulative frequency of vaccines of different types during follow up

**Supplementary figure 2**: Adjusted hazard ratios for portal vein thrombosis, other venous events, and death after ChAdOx1-S or BNT162b2 vaccine

**Supplementary Figure 3** Hazard ratios for major (A) arterial and (B) venous thrombotic events, and (C) haematological events, other events and lower limb fractures recorded in any position in EHR

**Supplementary Figure 4**: Hazard ratios for major (A) arterial and (B) venous thrombotic events, (C) haematological events, other events and fractures recorded in death record or in hospital record in first position followed by death <28 days

## SUPPLEMENTARY TABLES AND FIGURES

**Supplementary Table 1:** Hazard ratios (95% CIs) for thrombotic and other outcomes 1-28 and >28 days post-vaccination, compared with pre-vaccination rates. All analyses were stratified on geographical region (A) ChAdOx1-S vaccine (B) BNT162b2 vaccine

| Outcome | Age | Sex | 1-28 days post-vaccination |  |  | >28 days post vaccination s |  |  |
| --- | --- | --- | --- | --- | --- | --- | --- | --- |
|  |  |  | Unadjusted<sup>1</sup> | Age and sex adjusted<sup>2</sup> | Fully adjusted<sup>3</sup> | Unadjusted<sup>1</sup> | Age and sex adjusted<sup>2</sup> | Fully adjusted<sup>3</sup> |
| **(A) ChAdOx1-S vaccine**<br>**All venous events** | <70 | Overall | 2.33 (2.16–2.51) | 1.32 (1.22–1.43) | 0.97 (0.90–1.05) | 2.74 (2.39–3.14) | 1.60 (1.38–1.84) | 0.94 (0.81–1.08) |
|  |  | Men | 2.68 (2.41–2.97) | 1.33 (1.19–1.49) | 1.04 (0.93–1.16) | 3.13 (2.54–3.84) | 1.52 (1.23–1.89) | 0.97 (0.78–1.20) |
|  |  | Women | 2.08 (1.87–2.31) | 1.31 (1.17–1.47) | 0.96 (0.85–1.07) | 2.56 (2.13–3.07) | 1.67 (1.38–2.02) | 1.02 (0.84–1.23) |
|  | ≥70 | Overall | 0.65 (0.59–0.72) | 0.64 (0.59–0.70) | 0.58 (0.53–0.63) | 0.67 (0.59–0.76) | 0.58 (0.52–0.65) | 0.49 (0.44–0.55) |
|  |  | Men | 0.65 (0.56–0.76) | 0.65 (0.56–0.74) | 0.59 (0.51–0.68) | 0.70 (0.58–0.84) | 0.63 (0.53–0.75) | 0.55 (0.46–0.65) |
|  |  | Women | 0.65 (0.58–0.74) | 0.64 (0.56–0.71) | 0.57 (0.50–0.64) | 0.64 (0.54–0.76) | 0.54 (0.47–0.64) | 0.45 (0.39–0.53) |
| Intracranial venous thrombosis | <70 | Overall | 2.81 (1.70–4.64) | 2.99 (1.75–5.09) | 2.27 (1.33–3.88) | 2.37 (0.91–6.12) | 2.47 (0.95–6.42) | 1.72 (0.66–4.49) |
|  |  | Men | 2.72 (1.15–6.39) | 2.58 (0.99–6.74) | 2.06 (0.79–5.37) | 3.43 (0.74–15.87) | 3.37 (0.71–15.90) | 2.40 (0.53–10.86) |
|  |  | Women | 2.69 (1.45–5.00) | 3.27 (1.74–6.17) | 2.24 (1.17–4.31) | 1.75 (0.52–5.93) | 2.14 (0.63–7.22) | 1.47 (0.43–5.04) |
|  | ≥70 | Overall | 0.62 (0.17–2.23) | 0.65 (0.19–2.21) | 0.67 (0.20–2.18) | 1.35 (0.13–13.46) | 1.13 (0.14–9.20) | 1.11 (0.14–9.02) |
|  |  | Men | 10.8 (3.3–35.8) | 8.99 (2.75–29.42) | 10.1 (2.7–38.5) | - | - | - |
|  |  | Women | 0.27 (0.05–1.42) | 0.29 (0.06–1.43) | 0.28 (0.06–1.34) | 0.87 (0.10–7.32) | 0.82 (0.11–6.24) | 0.76 (0.10–5.70) |
| Portal vein thrombosis | <70 | Overall | 2.49 (1.20–5.14) | 1.56 (0.72–3.40) | 1.00 (0.24–4.15) | 2.76 (0.73–10.51) | 1.99 (0.53–7.52) | 1.00 (0.06–18.18) |
|  |  | Men | 3.38 (1.40–8.17) | 1.95 (0.76–5.02) | 1.22 (0.48–3.08) | 3.55 (0.52–24.13) | 2.16 (0.32–14.52) | 0.98 (0.14–6.75) |
|  |  | Women | 1.74 (0.48–6.26) | 1.06 (0.28–4.07) | 0.72 (0.18–2.86) | 2.39 (0.37–15.53) | 1.57 (0.24–10.12) | 0.86 (0.12–6.28) |
|  | ≥70 | Overall | 0.66 (0.11–4.06) | 0.67 (0.10–4.24) | 0.61 (0.10–3.71) | 2.72 (0.49–15.02) | 3.00 (0.48–18.99) | 2.61 (0.41–16.52) |
|  |  | Men | 0.43 (0.09–2.09) | 0.42 (0.08–2.15) | 0.37 (0.07–1.90) | 8.60 (2.33–31.70) | 7.99 (1.93–33.03) | 7.99 (2.03–31.38) |
|  |  | Women | 0.92 (0.06–13.23) | 0.92 (0.06–13.99) | 0.82 (0.06–11.05) | 0.87 (0.12–6.02) | 0.95 (0.12–7.39) | 0.86 (0.11–6.92) |
| Pulmonary embolism | <70 | Overall | 2.54 (2.30–2.80) | 1.28 (1.15–1.43) | 0.95 (0.85–1.05) | 3.12 (2.61–3.73) | 1.63 (1.34–1.96) | 0.95 (0.79–1.15) |
|  |  | Men | 2.83 (2.46–3.24) | 1.27 (1.09–1.48) | 1.01 (0.87–1.17) | 3.32 (2.52–4.38) | 1.43 (1.07–1.91) | 0.92 (0.69–1.23) |
|  |  | Women | 2.35 (2.04–2.70) | 1.30 (1.12–1.52) | 0.95 (0.82–1.10) | 3.09 (2.44–3.92) | 1.81 (1.41–2.32) | 1.09 (0.85–1.41) |
|  | ≥70 | Overall | 0.62 (0.55–0.70) | 0.60 (0.54–0.68) | 0.54 (0.48–0.61) | 0.61 (0.52–0.72) | 0.54 (0.46–0.62) | 0.45 (0.39–0.53) |
|  |  | Men | 0.63 (0.52–0.75) | 0.62 (0.52–0.74) | 0.57 (0.47–0.67) | 0.63 (0.50–0.79) | 0.57 (0.46–0.70) | 0.49 (0.40–0.62) |
|  |  | Women | 0.61 (0.52–0.72) | 0.59 (0.51–0.69) | 0.53 (0.45–0.61) | 0.60 (0.48–0.75) | 0.51 (0.42–0.63) | 0.43 (0.35–0.53) |
| Deep vein thrombosis | <70 | Overall | 2.08 (1.85–2.34) | 1.34 (1.18–1.52) | 0.99 (0.87–1.12) | 2.37 (1.90–2.96) | 1.58 (1.25–1.99) | 0.97 (0.77–1.22) |
|  |  | Men | 2.61 (2.22–3.07) | 1.48 (1.25–1.76) | 1.15 (0.97–1.36) | 2.85 (2.04–3.98) | 1.62 (1.14–2.29) | 1.06 (0.75–1.50) |
|  |  | Women | 1.67 (1.40–2.00) | 1.21 (1.00–1.46) | 0.90 (0.75–1.09) | 2.12 (1.57–2.86) | 1.57 (1.15–2.13) | 1.02 (0.75–1.40) |
|  | ≥70 | Overall | 0.73 (0.62–0.86) | 0.71 (0.61–0.83) | 0.63 (0.54–0.74) | 0.71 (0.57–0.88) | 0.62 (0.51–0.75) | 0.51 (0.42–0.62) |
|  |  | Men | 0.72 (0.56–0.94) | 0.71 (0.55–0.91) | 0.63 (0.49–0.81) | 0.77 (0.56–1.06) | 0.70 (0.51–0.95) | 0.58 (0.42–0.79) |
|  |  | Women | 0.73 (0.59–0.90) | 0.72 (0.59–0.87) | 0.64 (0.53–0.77) | 0.67 (0.51–0.88) | 0.57 (0.45–0.73) | 0.47 (0.37–0.60) |
| Other | <70 | Overall | 2.48 (1.70–3.62) | 1.60 (1.06–2.40) | 1.02 (0.69–1.52) | 3.04 (1.54–6.00) | 2.04 (1.01–4.09) | 0.94 (0.47–1.91) |
|  |  | Men | 2.61 (1.51–4.53) | 1.45 (0.79–2.64) | 1.02 (0.57–1.83) | 3.17 (1.13–8.90) | 1.74 (0.60–5.05) | 0.89 (0.31–2.58) |
|  |  | Women | 2.38 (1.42–4.00) | 1.74 (1.01–2.98) | 1.12 (0.65–1.91) | 3.15 (1.29–7.73) | 2.40 (0.97–5.96) | 1.26 (0.50–3.17) |
|  | ≥70 | Overall | 0.46 (0.21–1.01) | 0.46 (0.21–0.99) | 0.43 (0.20–0.92) | 1.13 (0.50–2.54) | 1.03 (0.48–2.21) | 0.93 (0.42–2.03) |
|  |  | Men | 0.38 (0.11–1.30) | 0.39 (0.12–1.31) | 0.33 (0.10–1.08) | 1.10 (0.36–3.39) | 1.02 (0.34–3.04) | 0.79 (0.26–2.36) |
|  |  | Women | 0.52 (0.19–1.44) | 0.51 (0.20–1.36) | 0.50 (0.19–1.32) | 1.16 (0.37–3.63) | 1.03 (0.36–2.98) | 0.98 (0.33–2.94) |
|  |  |  | Unadjusted<sup>1</sup> | Age and sex adjusted<sup>2</sup> | Fully adjusted<sup>3</sup> | Unadjusted<sup>1</sup> | Age and sex adjusted<sup>2</sup> | Fully adjusted<sup>3</sup> |
| All arterial events | <70 | Overall | 3.02 (2.90–3.14) | 1.25 (1.20–1.31) | 0.90 (0.86–0.95) | 3.55 (3.29–3.83) | 1.57 (1.45–1.70) | 0.91 (0.84–1.00) |
|  |  | Men | 3.42 (3.26–3.60) | 1.26 (1.20–1.34) | 0.91 (0.86–0.96) | 4.70 (4.27–5.17) | 1.66 (1.50–1.84) | 0.96 (0.86–1.07) |
|  |  | Women | 2.96 (2.76–3.18) | 1.24 (1.14–1.34) | 0.93 (0.86–1.00) | 3.30 (2.90–3.75) | 1.43 (1.25–1.64) | 0.88 (0.77–1.02) |
|  | ≥70 | Overall | 0.89 (0.85–0.93) | 0.84 (0.80–0.87) | 0.76 (0.73–0.79) | 1.03 (0.97–1.10) | 0.83 (0.78–0.87) | 0.72 (0.68–0.77) |
|  |  | Men | 0.90 (0.85–0.96) | 0.87 (0.82–0.92) | 0.78 (0.74–0.83) | 1.06 (0.97–1.16) | 0.89 (0.82–0.96) | 0.78 (0.71–0.85) |
|  |  | Women | 0.88 (0.82–0.94) | 0.80 (0.76–0.85) | 0.73 (0.69–0.78) | 1.02 (0.93–1.12) | 0.77 (0.71–0.83) | 0.68 (0.62–0.74) |
| Myocardial infarction | <70 | Overall | 2.85 (2.70–3.01) | 1.24 (1.17–1.32) | 0.88 (0.83–0.94) | 2.93 (2.63–3.26) | 1.40 (1.24–1.56) | 0.83 (0.73–0.93) |
|  |  | Men | 3.23 (3.02–3.44) | 1.25 (1.17–1.35) | 0.89 (0.83–0.95) | 3.52 (3.08–4.03) | 1.33 (1.15–1.53) | 0.78 (0.68–0.90) |
|  |  | Women | 2.93 (2.63–3.26) | 1.22 (1.09–1.38) | 0.92 (0.82–1.04) | 3.56 (2.95–4.29) | 1.58 (1.29–1.92) | 1.01 (0.82–1.24) |
|  | ≥70 | Overall | 0.86 (0.80–0.92) | 0.83 (0.78–0.88) | 0.76 (0.71–0.81) | 0.97 (0.89–1.07) | 0.83 (0.76–0.90) | 0.74 (0.68–0.81) |
|  |  | Men | 0.88 (0.80–0.96) | 0.85 (0.78–0.93) | 0.77 (0.71–0.84) | 0.96 (0.84–1.09) | 0.84 (0.74–0.94) | 0.73 (0.64–0.82) |
|  |  | Women | 0.85 (0.76–0.94) | 0.80 (0.72–0.88) | 0.76 (0.69–0.84) | 1.03 (0.90–1.19) | 0.82 (0.72–0.93) | 0.78 (0.68–0.88) |
| Ischaemic stroke<sup>4</sup> | <70 | Overall | 3.21 (3.02–3.41) | 1.25 (1.17–1.34) | 0.90 (0.84–0.96) | 4.36 (3.91–4.86) | 1.76 (1.56–1.98) | 0.94 (0.84–1.07) |
|  |  | Men | 3.73 (3.44–4.05) | 1.28 (1.17–1.40) | 0.91 (0.83–0.99) | 6.58 (5.73–7.57) | 2.14 (1.84–2.49) | 1.12 (0.96–1.31) |
|  |  | Women | 2.91 (2.64–3.20) | 1.21 (1.09–1.35) | 0.90 (0.81–1.00) | 3.10 (2.59–3.71) | 1.32 (1.09–1.59) | 0.77 (0.63–0.94) |
|  | ≥70 | Overall | 0.92 (0.87–0.98) | 0.85 (0.81–0.90) | 0.77 (0.73–0.82) | 1.08 (0.99–1.18) | 0.83 (0.77–0.90) | 0.72 (0.67–0.78) |
|  |  | Men | 0.94 (0.85–1.03) | 0.89 (0.82–0.96) | 0.81 (0.74–0.88) | 1.14 (1.01–1.30) | 0.93 (0.83–1.04) | 0.82 (0.73–0.92) |
|  |  | Women | 0.92 (0.84–1.00) | 0.82 (0.77–0.89) | 0.75 (0.69–0.80) | 1.04 (0.92–1.17) | 0.76 (0.68–0.84) | 0.66 (0.59–0.73) |
| Other arterial | <70 | Overall | 3.18 (2.48–4.07) | 1.24 (0.93–1.64) | 0.82 (0.63–1.07) | 4.20 (2.62–6.73) | 1.68 (0.99–2.86) | 0.77 (0.46–1.30) |
|  |  | Men | 3.17 (2.29–4.40) | 1.05 (0.73–1.51) | 0.75 (0.53–1.07) | 5.56 (3.07–10.06) | 1.66 (0.86–3.20) | 0.89 (0.47–1.69) |
|  |  | Women | 3.67 (2.51–5.37) | 1.59 (1.03–2.46) | 1.04 (0.68–1.59) | 3.60 (1.64–7.91) | 1.74 (0.73–4.18) | 0.83 (0.35–2.01) |
|  | ≥70 | Overall | 0.44 (0.33–0.59) | 0.45 (0.34–0.59) | 0.41 (0.31–0.53) | 0.55 (0.38–0.79) | 0.49 (0.35–0.70) | 0.44 (0.31–0.63) |
|  |  | Men | 0.46 (0.31–0.67) | 0.45 (0.31–0.66) | 0.42 (0.29–0.61) | 0.61 (0.35–1.05) | 0.56 (0.33–0.95) | 0.51 (0.30–0.87) |
|  |  | Women | 0.43 (0.28–0.66) | 0.44 (0.30–0.67) | 0.39 (0.26–0.59) | 0.51 (0.30–0.86) | 0.44 (0.27–0.71) | 0.38 (0.23–0.62) |
| Haematological events |  |  |  |  |  |  |  |  |
| Any thrombocytopenia | <70 | Overall | 4.85 (3.87–6.07) | 3.00 (2.34–3.85) | 1.71 (1.35–2.16) | 7.48 (5.31–10.53) | 4.68 (3.24–6.77) | 1.69 (1.16–2.46) |
|  |  | Men | 6.47 (4.62–9.06) | 3.52 (2.42–5.13) | 1.85 (1.30–2.63) | 13.2 (8.1–21.4) | 6.84 (3.99–11.73) | 2.02 (1.18–3.44) |
|  |  | Women | 3.81 (2.80–5.17) | 2.65 (1.89–3.71) | 1.95 (1.40–2.72) | 4.59 (2.78–7.58) | 3.34 (1.98–5.64) | 2.08 (1.22–3.54) |
|  | ≥70 | Overall | 0.92 (0.65–1.29) | 0.87 (0.63–1.22) | 0.79 (0.56–1.10) | 1.14 (0.66–1.95) | 0.99 (0.59–1.65) | 0.84 (0.50–1.41) |
|  |  | Men | 0.84 (0.53–1.32) | 0.79 (0.50–1.22) | 0.68 (0.43–1.06) | 1.37 (0.64–2.91) | 1.17 (0.57–2.38) | 0.92 (0.45–1.90) |
|  |  | Women | 1.04 (0.62–1.75) | 0.99 (0.61–1.63) | 0.93 (0.56–1.54) | 1.02 (0.46–2.24) | 0.88 (0.42–1.85) | 0.80 (0.38–1.70) |
| Other events |  |  |  |  |  |  |  |  |
| Haemorrhagic stroke<sup>5</sup> | <70 | Overall | 2.25 (1.88–2.69) | 1.05 (0.86–1.27) | 1.00 (0.73–1.37) | 2.87 (2.10–3.92) | 1.39 (1.00–1.93) | 1.00 (0.51–1.95) |
|  |  | Men | 2.68 (2.11–3.41) | 1.15 (0.89–1.49) | 1.01 (0.78–1.30) | 2.86 (1.74–4.71) | 1.21 (0.72–2.04) | 0.87 (0.52–1.46) |
|  |  | Women | 1.93 (1.48–2.52) | 0.93 (0.70–1.24) | 0.87 (0.66–1.16) | 2.94 (1.95–4.42) | 1.47 (0.95–2.27) | 1.23 (0.80–1.91) |
|  | ≥70 | Overall | 0.83 (0.69–1.00) | 0.78 (0.66–0.92) | 0.73 (0.62–0.87) | 1.09 (0.84–1.40) | 0.84 (0.67–1.04) | 0.76 (0.61–0.95) |
|  |  | Men | 0.86 (0.65–1.14) | 0.82 (0.64–1.05) | 0.79 (0.62–1.01) | 1.34 (0.93–1.94) | 1.03 (0.75–1.43) | 0.97 (0.70–1.34) |
|  |  | Women | 0.81 (0.63–1.03) | 0.75 (0.60–0.94) | 0.70 (0.56–0.87) | 0.92 (0.65–1.30) | 0.71 (0.52–0.96) | 0.64 (0.47–0.86) |
|  |  |  | Unadjusted<sup>1</sup> | Age and sex adjusted<sup>2</sup> | Fully adjusted<sup>3</sup> | Unadjusted<sup>1</sup> | Age and sex adjusted<sup>2</sup> | Fully adjusted<sup>3</sup> |
| Mesenteric thrombosis | <70 | Overall | 3.25 (2.42–4.37) | 1.24 (0.89–1.73) | 0.84 (0.61–1.16) | 5.47 (3.33–8.99) | 2.16 (1.24–3.77) | 1.04 (0.60–1.80) |
|  |  | Men | 2.87 (1.72–4.77) | 0.98 (0.57–1.71) | 0.72 (0.43–1.22) | 4.34 (1.59–11.83) | 1.42 (0.48–4.25) | 0.75 (0.26–2.17) |
|  |  | Women | 3.39 (2.36–4.87) | 1.43 (0.94–2.17) | 1.01 (0.67–1.52) | 5.66 (3.21–9.99) | 2.59 (1.36–4.94) | 1.40 (0.73–2.69) |
|  | ≥70 | Overall | 0.66 (0.49–0.89) | 0.60 (0.46–0.79) | 0.53 (0.41–0.70) | 0.95 (0.64–1.43) | 0.74 (0.52–1.05) | 0.62 (0.43–0.89) |
|  |  | Men | 0.65 (0.39–1.09) | 0.61 (0.38–0.98) | 0.52 (0.32–0.83) | 0.81 (0.44–1.49) | 0.69 (0.40–1.18) | 0.54 (0.31–0.94) |
|  |  | Women | 0.66 (0.46–0.95) | 0.60 (0.44–0.84) | 0.55 (0.40–0.77) | 1.00 (0.60–1.68) | 0.76 (0.48–1.20) | 0.67 (0.42–1.08) |
| Lower limb fracture | <70 | Overall | 1.71 (1.55–1.87) | 1.01 (0.91–1.11) | 0.85 (0.77–0.93) | 2.26 (1.91–2.67) | 1.35 (1.13–1.60) | 0.98 (0.83–1.17) |
|  |  | Men | 1.35 (1.15–1.58) | 0.98 (0.83–1.16) | 0.86 (0.73–1.02) | 2.22 (1.67–2.93) | 1.60 (1.20–2.14) | 1.24 (0.93–1.66) |
|  |  | Women | 1.91 (1.70–2.15) | 1.03 (0.91–1.16) | 0.88 (0.78–0.99) | 2.21 (1.80–2.72) | 1.25 (1.01–1.55) | 0.95 (0.77–1.18) |
|  | ≥70 | Overall | 1.11 (1.02–1.21) | 0.94 (0.88–1.01) | 0.83 (0.78–0.89) | 1.56 (1.39–1.76) | 0.95 (0.87–1.04) | 0.78 (0.71–0.85) |
|  |  | Men | 1.06 (0.90–1.24) | 0.94 (0.83–1.08) | 0.84 (0.73–0.95) | 1.50 (1.19–1.89) | 0.97 (0.81–1.17) | 0.81 (0.67–0.97) |
|  |  | Women | 1.11 (1.01–1.22) | 0.95 (0.88–1.03) | 0.84 (0.77–0.91) | 1.51 (1.32–1.73) | 0.95 (0.85–1.06) | 0.78 (0.70–0.87) |
| Death | <70 | Overall | 1.96 (1.87–2.06) | 0.60 (0.57–0.63) | 0.37 (0.35–0.39) | 4.51 (4.20–4.85) | 1.41 (1.30–1.53) | 0.51 (0.47–0.56) |
|  |  | Men | 2.35 (2.20–2.51) | 0.63 (0.58–0.67) | 0.44 (0.41–0.47) | 6.47 (5.87–7.13) | 1.57 (1.40–1.75) | 0.75 (0.67–0.85) |
|  |  | Women | 1.69 (1.56–1.82) | 0.56 (0.52–0.61) | 0.36 (0.33–0.39) | 3.48 (3.13–3.89) | 1.21 (1.07–1.37) | 0.50 (0.44–0.57) |
|  | ≥70 | Overall | 0.35 (0.34–0.36) | 0.34 (0.33–0.35) | 0.28 (0.28–0.29) | 0.49 (0.47–0.51) | 0.38 (0.37–0.39) | 0.27 (0.26–0.28) |
|  |  | Men | 0.31 (0.30–0.32) | 0.32 (0.31–0.33) | 0.26 (0.25–0.27) | 0.44 (0.42–0.47) | 0.37 (0.35–0.38) | 0.26 (0.24–0.27) |
|  |  | Women | 0.39 (0.37–0.40) | 0.36 (0.35–0.37) | 0.30 (0.29–0.31) | 0.54 (0.51–0.56) | 0.39 (0.38–0.41) | 0.29 (0.27–0.30) |
|  |  |  | Unadjusted<sup>1</sup> | Age and sex adjusted<sup>2</sup> | Fully adjusted<sup>3</sup> | Unadjusted<sup>1</sup> | Age and sex adjusted<sup>2</sup> | Fully adjusted<sup>3</sup> |
| **(B) BNT162b2 vaccine**<br>**All venous events** | <70 | Overall | 1.47 (1.35–1.61) | 1.01 (0.93–1.11) | 0.81 (0.74–0.88) | 1.14 (1.01–1.29) | 0.88 (0.77–1.00) | 0.73 (0.64–0.83) |
|  |  | Men | 1.98 (1.75–2.23) | 1.15 (1.01–1.30) | 0.92 (0.81–1.05) | 1.56 (1.28–1.89) | 1.02 (0.83–1.25) | 0.83 (0.68–1.02) |
|  |  | Women | 1.20 (1.06–1.37) | 0.91 (0.80–1.03) | 0.74 (0.65–0.84) | 1.01 (0.86–1.20) | 0.81 (0.69–0.96) | 0.70 (0.59–0.83) |
|  | ≥70 | Overall | 0.65 (0.60–0.71) | 0.56 (0.52–0.61) | 0.57 (0.53–0.62) | 0.68 (0.62–0.75) | 0.50 (0.46–0.55) | 0.50 (0.45–0.55) |
|  |  | Men | 0.67 (0.59–0.76) | 0.59 (0.52–0.66) | 0.60 (0.53–0.67) | 0.75 (0.65–0.87) | 0.59 (0.51–0.68) | 0.58 (0.50–0.67) |
|  |  | Women | 0.64 (0.58–0.72) | 0.54 (0.49–0.60) | 0.56 (0.50–0.63) | 0.63 (0.56–0.72) | 0.45 (0.40–0.51) | 0.45 (0.40–0.52) |
| Intracranial venous thrombosis | <70 | Overall | 0.76 (0.31–1.85) | 0.72 (0.29–1.78) | 0.59 (0.24–1.45) | 0.65 (0.20–2.13) | 0.59 (0.18–1.93) | 0.51 (0.16–1.68) |
|  |  | Men | 1.60 (0.50–5.16) | 1.39 (0.43–4.49) | 1.09 (0.33–3.61) | 1.04 (0.14–7.77) | 0.92 (0.12–7.05) | 0.74 (0.10–5.62) |
|  |  | Women | 0.39 (0.10–1.60) | 0.44 (0.11–1.78) | 0.37 (0.09–1.52) | 0.48 (0.11–2.07) | 0.51 (0.12–2.20) | 0.45 (0.10–1.95) |
|  | ≥70 | Overall | 1.65 (0.64–4.29) | 1.25 (0.48–3.26) | 1.43 (0.55–3.75) | 1.13 (0.13–9.75) | 0.71 (0.08–6.56) | 0.88 (0.10–7.52) |
|  |  | Men | - | - | - | - | - | - |
|  |  | Women | 2.01 (0.69–5.88) | 1.45 (0.49–4.26) | 1.57 (0.54–4.54) | 1.26 (0.13–11.94) | 0.73 (0.07–7.29) | 0.82 (0.09–7.61) |
| Portal vein thrombosis | <70 | Overall | 0.58 (0.15–2.30) | 0.44 (0.11–1.72) | 0.29 (0.07–1.17) | 3.06 (1.29–7.24) | 2.84 (1.19–6.74) | 2.19 (0.89–5.37) |
|  |  | Men | 1.28 (0.33–4.95) | 0.81 (0.21–3.09) | 0.51 (0.13–2.04) | - | - | - |
|  |  | Women | - | - | - | 5.30 (1.98–14.22) | 4.28 (1.55–11.78) | 3.95 (1.39–11.20) |
|  | ≥70 | Overall | 0.49 (0.10–2.28) | 0.43 (0.10–1.87) | 0.42 (0.09–1.91) | 0.67 (0.23–1.97) | 0.54 (0.19–1.56) | 0.48 (0.17–1.38) |
|  |  | Men | 0.85 (0.17–4.38) | 0.55 (0.11–2.66) | 0.49 (0.10–2.46) | 1.26 (0.36–4.41) | 0.70 (0.23–2.17) | 0.48 (0.17–1.38) |
|  |  | Women | 0.22 (0.01–3.38) | 0.24 (0.01–3.80) | 0.26 (0.02–4.47) | 0.37 (0.09–1.52) | 0.44 (0.10–2.01) | 0.46 (0.11–2.04) |
| Pulmonary embolism | <70 | Overall | 1.52 (1.36–1.71) | 0.99 (0.87–1.11) | 0.78 (0.69–0.88) | 1.20 (1.02–1.43) | 0.89 (0.75–1.06) | 0.71 (0.60–0.85) |
|  |  | Men | 1.93 (1.63–2.28) | 1.05 (0.88–1.25) | 0.85 (0.71–1.01) | 1.56 (1.19–2.05) | 0.97 (0.74–1.29) | 0.77 (0.58–1.03) |
|  |  | Women | 1.32 (1.13–1.56) | 0.94 (0.79–1.10) | 0.75 (0.64–0.89) | 1.10 (0.88–1.37) | 0.84 (0.67–1.05) | 0.71 (0.57–0.89) |
|  | ≥70 | Overall | 0.61 (0.55–0.68) | 0.53 (0.48–0.59) | 0.54 (0.49–0.60) | 0.61 (0.54–0.69) | 0.45 (0.40–0.51) | 0.45 (0.39–0.50) |
|  |  | Men | 0.64 (0.55–0.75) | 0.56 (0.48–0.65) | 0.57 (0.49–0.66) | 0.65 (0.54–0.77) | 0.50 (0.42–0.59) | 0.49 (0.41–0.59) |
|  |  | Women | 0.59 (0.52–0.68) | 0.51 (0.44–0.58) | 0.52 (0.45–0.60) | 0.58 (0.49–0.68) | 0.42 (0.36–0.49) | 0.42 (0.36–0.49) |
| Deep vein thrombosis | <70 | Overall | 1.38 (1.21–1.59) | 1.03 (0.89–1.18) | 0.82 (0.71–0.95) | 1.05 (0.86–1.28) | 0.85 (0.69–1.04) | 0.73 (0.59–0.89) |
|  |  | Men | 1.93 (1.60–2.33) | 1.23 (1.01–1.49) | 0.99 (0.81–1.20) | 1.55 (1.15–2.10) | 1.10 (0.81–1.48) | 0.94 (0.69–1.28) |
|  |  | Women | 1.08 (0.88–1.33) | 0.87 (0.71–1.08) | 0.72 (0.58–0.89) | 0.88 (0.67–1.15) | 0.74 (0.56–0.97) | 0.65 (0.50–0.86) |
|  | ≥70 | Overall | 0.69 (0.60–0.79) | 0.58 (0.51–0.67) | 0.61 (0.53–0.70) | 0.81 (0.69–0.96) | 0.59 (0.50–0.70) | 0.61 (0.52–0.73) |
|  |  | Men | 0.66 (0.53–0.82) | 0.59 (0.48–0.73) | 0.61 (0.49–0.76) | 0.95 (0.74–1.23) | 0.78 (0.60–1.01) | 0.80 (0.61–1.05) |
|  |  | Women | 0.71 (0.59–0.85) | 0.59 (0.49–0.70) | 0.62 (0.52–0.74) | 0.72 (0.58–0.90) | 0.49 (0.40–0.61) | 0.52 (0.41–0.64) |
| Other | <70 | Overall | 2.16 (1.47–3.19) | 1.60 (1.07–2.37) | 1.18 (0.79–1.77) | 1.64 (0.93–2.90) | 1.34 (0.75–2.40) | 1.03 (0.57–1.87) |
|  |  | Men | 3.25 (1.93–5.50) | 2.03 (1.16–3.54) | 1.46 (0.83–2.57) | 2.14 (0.86–5.34) | 1.51 (0.59–3.86) | 1.03 (0.39–2.70) |
|  |  | Women | 1.48 (0.82–2.65) | 1.20 (0.67–2.16) | 0.98 (0.54–1.77) | 1.55 (0.75–3.21) | 1.31 (0.63–2.74) | 1.21 (0.57–2.55) |
|  | ≥70 | Overall | 1.13 (0.66–1.94) | 0.97 (0.58–1.64) | 1.01 (0.60–1.72) | 0.86 (0.39–1.88) | 0.66 (0.31–1.41) | 0.67 (0.30–1.47) |
|  |  | Men | 1.50 (0.77–2.95) | 1.25 (0.65–2.42) | 1.27 (0.64–2.49) | 1.51 (0.50–4.51) | 1.09 (0.36–3.34) | 1.07 (0.33–3.44) |
|  |  | Women | 0.79 (0.31–2.04) | 0.72 (0.30–1.75) | 0.75 (0.30–1.85) | 0.42 (0.17–1.06) | 0.36 (0.15–0.88) | 0.36 (0.14–0.91) |
| **All arterial events** | <70 | Overall | 2.21 (2.12–2.31) | 1.31 (1.25–1.37) | 0.94 (0.90–0.99) | 1.45 (1.35–1.55) | 1.08 (1.01–1.16) | 0.88 (0.82–0.95) |
|  |  | Men | 3.06 (2.90–3.22) | 1.44 (1.36–1.52) | 0.99 (0.93–1.05) | 2.17 (1.99–2.38) | 1.19 (1.08–1.31) | 0.91 (0.82–1.00) |
|  |  | Women | 1.85 (1.71–1.99) | 1.11 (1.02–1.20) | 0.92 (0.84–0.99) | 1.44 (1.30–1.60) | 0.96 (0.86–1.07) | 0.91 (0.81–1.02) |
|  | ≥70 | Overall | 0.90 (0.87–0.93) | 0.69 (0.66–0.71) | 0.72 (0.70–0.75) | 1.15 (1.10–1.20) | 0.69 (0.66–0.72) | 0.71 (0.68–0.74) |
|  |  | Men | 0.96 (0.91–1.00) | 0.75 (0.71–0.78) | 0.77 (0.73–0.81) | 1.17 (1.11–1.25) | 0.74 (0.70–0.79) | 0.74 (0.70–0.79) |
|  |  | Women | 0.84 (0.80–0.89) | 0.63 (0.60–0.66) | 0.68 (0.65–0.71) | 1.13 (1.06–1.20) | 0.64 (0.61–0.68) | 0.68 (0.64–0.73) |
|  |  |  | Unadjusted<sup>1</sup> | Age and sex adjusted<sup>2</sup> | Fully adjusted<sup>3</sup> | Unadjusted<sup>1</sup> | Age and sex adjusted<sup>2</sup> | Fully adjusted<sup>3</sup> |
| Myocardial infarction | <70 | Overall | 2.24 (2.12–2.37) | 1.39 (1.30–1.47) | 0.94 (0.88–1.00) | 1.42 (1.29–1.55) | 1.14 (1.04–1.26) | 0.88 (0.80–0.97) |
|  |  | Men | 3.07 (2.87–3.28) | 1.51 (1.40–1.62) | 0.97 (0.90–1.05) | 1.98 (1.76–2.23) | 1.13 (1.00–1.28) | 0.81 (0.71–0.93) |
|  |  | Women | 1.94 (1.74–2.17) | 1.15 (1.03–1.30) | 0.94 (0.83–1.06) | 1.78 (1.54–2.05) | 1.18 (1.02–1.37) | 1.11 (0.95–1.30) |
|  | ≥70 | Overall | 0.93 (0.88–0.98) | 0.74 (0.71–0.78) | 0.74 (0.70–0.78) | 1.19 (1.12–1.27) | 0.78 (0.73–0.83) | 0.75 (0.70–0.80) |
|  |  | Men | 0.99 (0.92–1.06) | 0.81 (0.76–0.86) | 0.79 (0.73–0.84) | 1.20 (1.10–1.31) | 0.82 (0.76–0.89) | 0.76 (0.70–0.83) |
|  |  | Women | 0.85 (0.78–0.92) | 0.66 (0.61–0.72) | 0.69 (0.63–0.74) | 1.20 (1.08–1.33) | 0.74 (0.67–0.81) | 0.74 (0.67–0.82) |
| Ischaemic stroke<sup>4</sup> | <70 | Overall | 2.18 (2.05–2.33) | 1.23 (1.14–1.31) | 0.90 (0.83–0.97) | 1.46 (1.32–1.62) | 1.00 (0.89–1.11) | 0.80 (0.72–0.90) |
|  |  | Men | 3.05 (2.80–3.32) | 1.38 (1.26–1.51) | 0.95 (0.86–1.05) | 2.40 (2.08–2.77) | 1.26 (1.09–1.46) | 0.91 (0.78–1.07) |
|  |  | Women | 1.76 (1.59–1.96) | 1.05 (0.94–1.17) | 0.85 (0.76–0.95) | 1.18 (1.01–1.38) | 0.77 (0.66–0.90) | 0.73 (0.61–0.86) |
|  | ≥70 | Overall | 0.88 (0.84–0.93) | 0.65 (0.63–0.68) | 0.71 (0.68–0.75) | 1.13 (1.06–1.19) | 0.64 (0.60–0.67) | 0.69 (0.66–0.73) |
|  |  | Men | 0.94 (0.88–1.00) | 0.71 (0.66–0.75) | 0.77 (0.72–0.82) | 1.17 (1.07–1.27) | 0.69 (0.64–0.75) | 0.75 (0.69–0.81) |
|  |  | Women | 0.84 (0.78–0.89) | 0.61 (0.58–0.65) | 0.68 (0.63–0.72) | 1.09 (1.01–1.19) | 0.60 (0.56–0.65) | 0.66 (0.61–0.71) |
| Other arterial | <70 | Overall | 1.72 (1.28–2.31) | 0.95 (0.70–1.30) | 0.69 (0.50–0.94) | 1.63 (1.10–2.43) | 1.13 (0.74–1.72) | 0.85 (0.55–1.32) |
|  |  | Men | 2.31 (1.58–3.37) | 0.99 (0.66–1.49) | 0.71 (0.47–1.07) | 3.22 (2.02–5.13) | 1.56 (0.95–2.58) | 1.16 (0.69–1.96) |
|  |  | Women | 1.46 (0.90–2.37) | 0.88 (0.54–1.45) | 0.69 (0.42–1.15) | 0.93 (0.43–1.99) | 0.65 (0.30–1.43) | 0.59 (0.26–1.33) |
|  | ≥70 | Overall | 0.59 (0.46–0.76) | 0.51 (0.40–0.65) | 0.52 (0.41–0.67) | 0.64 (0.47–0.85) | 0.47 (0.35–0.62) | 0.46 (0.34–0.62) |
|  |  | Men | 0.49 (0.35–0.69) | 0.46 (0.32–0.65) | 0.47 (0.33–0.66) | 0.52 (0.37–0.74) | 0.46 (0.32–0.66) | 0.44 (0.31–0.64) |
|  |  | Women | 0.74 (0.51–1.08) | 0.59 (0.41–0.84) | 0.60 (0.42–0.85) | 0.83 (0.50–1.37) | 0.50 (0.31–0.79) | 0.49 (0.30–0.78) |
| Haematological events |  |  |  |  |  |  |  |  |
| Any thrombocytopenia | <70 | Overall | 2.10 (1.60–2.77) | 1.48 (1.11–1.97) | 1.00 (0.75–1.34) | 2.03 (1.42–2.92) | 1.54 (1.07–2.23) | 0.97 (0.66–1.41) |
|  |  | Men | 3.31 (2.23–4.91) | 2.07 (1.35–3.16) | 1.19 (0.78–1.83) | 2.95 (1.61–5.42) | 2.02 (1.08–3.79) | 0.93 (0.49–1.78) |
|  |  | Women | 1.50 (1.01–2.22) | 1.17 (0.78–1.74) | 0.97 (0.65–1.46) | 1.58 (1.00–2.50) | 1.32 (0.83–2.08) | 1.14 (0.72–1.80) |
|  | ≥70 | Overall | 0.85 (0.64–1.13) | 0.72 (0.54–0.95) | 0.68 (0.51–0.90) | 1.22 (0.87–1.71) | 0.90 (0.64–1.25) | 0.79 (0.56–1.12) |
|  |  | Men | 0.87 (0.61–1.26) | 0.73 (0.51–1.05) | 0.68 (0.47–0.97) | 1.55 (1.03–2.32) | 1.12 (0.75–1.67) | 0.95 (0.62–1.46) |
|  |  | Women | 0.82 (0.53–1.29) | 0.71 (0.46–1.09) | 0.68 (0.44–1.07) | 0.84 (0.48–1.48) | 0.63 (0.36–1.09) | 0.57 (0.32–1.02) |
| Other events |  |  |  |  |  |  |  |  |
| Haemorrhagic stroke<sup>5</sup> | <70 | Overall | 1.39 (1.13–1.71) | 0.83 (0.67–1.03) | 0.77 (0.62–0.96) | 1.31 (1.00–1.73) | 0.90 (0.68–1.19) | 0.86 (0.65–1.15) |
|  |  | Men | 1.65 (1.22–2.23) | 0.86 (0.63–1.18) | 0.77 (0.57–1.05) | 1.43 (0.89–2.29) | 0.86 (0.53–1.40) | 0.77 (0.47–1.25) |
|  |  | Women | 1.28 (0.95–1.72) | 0.80 (0.59–1.08) | 0.78 (0.58–1.05) | 1.34 (0.95–1.88) | 0.89 (0.62–1.26) | 0.92 (0.65–1.31) |
|  | ≥70 | Overall | 0.80 (0.69–0.91) | 0.59 (0.52–0.68) | 0.65 (0.57–0.74) | 1.28 (1.09–1.50) | 0.72 (0.62–0.84) | 0.80 (0.68–0.94) |
|  |  | Men | 0.82 (0.67–1.01) | 0.59 (0.48–0.71) | 0.64 (0.52–0.78) | 1.45 (1.14–1.83) | 0.78 (0.62–0.98) | 0.86 (0.68–1.09) |
|  |  | Women | 0.78 (0.65–0.94) | 0.60 (0.50–0.72) | 0.66 (0.55–0.79) | 1.16 (0.93–1.44) | 0.68 (0.56–0.84) | 0.76 (0.61–0.93) |
| Mesenteric thrombosis | <70 | Overall | 1.61 (1.10–2.36) | 0.84 (0.56–1.26) | 0.65 (0.43–0.97) | 1.77 (1.11–2.84) | 1.08 (0.66–1.75) | 0.83 (0.50–1.37) |
|  |  | Men | 2.91 (1.78–4.75) | 1.29 (0.77–2.17) | 0.97 (0.57–1.64) | 2.21 (0.99–4.94) | 1.15 (0.50–2.62) | 0.82 (0.35–1.91) |
|  |  | Women | 0.91 (0.49–1.67) | 0.55 (0.29–1.03) | 0.45 (0.23–0.86) | 1.61 (0.90–2.88) | 1.09 (0.60–1.98) | 0.94 (0.50–1.75) |
|  | ≥70 | Overall | 0.72 (0.57–0.90) | 0.54 (0.43–0.67) | 0.54 (0.44–0.68) | 1.06 (0.81–1.37) | 0.61 (0.48–0.78) | 0.59 (0.45–0.76) |
|  |  | Men | 0.57 (0.37–0.86) | 0.41 (0.27–0.61) | 0.42 (0.28–0.62) | 0.82 (0.53–1.29) | 0.44 (0.29–0.66) | 0.42 (0.27–0.65) |
|  |  | Women | 0.80 (0.61–1.04) | 0.61 (0.47–0.80) | 0.62 (0.48–0.81) | 1.18 (0.86–1.63) | 0.72 (0.53–0.98) | 0.70 (0.51–0.96) |
| Lower limb fracture | <70 | Overall | 1.44 (1.31–1.59) | 1.03 (0.93–1.14) | 0.93 (0.84–1.02) | 1.24 (1.08–1.42) | 0.95 (0.83–1.09) | 0.89 (0.78–1.03) |
|  |  | Men | 1.19 (0.99–1.42) | 0.94 (0.78–1.12) | 0.85 (0.71–1.02) | 0.99 (0.74–1.31) | 0.83 (0.62–1.11) | 0.80 (0.60–1.07) |
|  |  | Women | 1.49 (1.33–1.67) | 1.08 (0.96–1.21) | 0.98 (0.87–1.10) | 1.27 (1.09–1.48) | 1.00 (0.85–1.17) | 0.94 (0.80–1.11) |
|  | ≥70 | Overall | 0.99 (0.93–1.05) | 0.65 (0.62–0.69) | 0.69 (0.66–0.73) | 1.48 (1.38–1.59) | 0.67 (0.63–0.72) | 0.71 (0.66–0.76) |
|  |  | Men | 0.95 (0.85–1.06) | 0.58 (0.52–0.64) | 0.63 (0.57–0.70) | 1.60 (1.40–1.83) | 0.65 (0.57–0.73) | 0.69 (0.61–0.79) |
|  |  | Women | 1.00 (0.93–1.07) | 0.68 (0.64–0.73) | 0.72 (0.67–0.77) | 1.42 (1.30–1.55) | 0.68 (0.63–0.73) | 0.71 (0.65–0.77) |
|  |  |  | Unadjusted<sup>1</sup> | Age and sex adjusted<sup>2</sup> | Fully adjusted<sup>3</sup> | Unadjusted<sup>1</sup> | Age and sex adjusted<sup>2</sup> | Fully adjusted<sup>3</sup> |
| Death | <70 | Overall | 0.74 (0.69–0.79) | 0.35 (0.32–0.37) | 0.24 (0.22–0.26) | 0.91 (0.84–1.00) | 0.52 (0.48–0.57) | 0.33 (0.30–0.37) |
|  |  | Men | 1.06 (0.97–1.17) | 0.39 (0.36–0.43) | 0.28 (0.25–0.31) | 1.62 (1.44–1.82) | 0.69 (0.61–0.79) | 0.44 (0.39–0.51) |
|  |  | Women | 0.56 (0.50–0.63) | 0.29 (0.26–0.33) | 0.22 (0.19–0.25) | 0.66 (0.58–0.75) | 0.38 (0.34–0.44) | 0.28 (0.25–0.33) |
|  | ≥70 | Overall | 0.22 (0.22–0.23) | 0.16 (0.16–0.16) | 0.19 (0.19–0.20) | 0.30 (0.29–0.31) | 0.16 (0.16–0.16) | 0.19 (0.18–0.19) |
|  |  | Men | 0.23 (0.22–0.24) | 0.16 (0.15–0.16) | 0.19 (0.18–0.20) | 0.33 (0.32–0.34) | 0.16 (0.15–0.17) | 0.19 (0.18–0.19) |
|  |  | Women | 0.21 (0.20–0.22) | 0.16 (0.16–0.17) | 0.20 (0.19–0.20) | 0.27 (0.26–0.28) | 0.16 (0.15–0.17) | 0.19 (0.18–0.19) |

<sup>1</sup>Stratified by region
<sup>2</sup>Adjusted for age, age<sup>2</sup>, sex and stratified by region
<sup>3</sup>Adjusted for age, age<sup>2</sup>, sex, ethnicity, deprivation, smoking and co-morbidities, and stratified by region
<sup>4</sup>Ischaemic stroke: including ischaemic stroke, stroke of uncertain cause, and spinal stroke
<sup>5</sup>Hemorrhagic stroke: including intracranial hemorrhage and subarachnoid hemorrhage

**Supplementary table 2(a).**
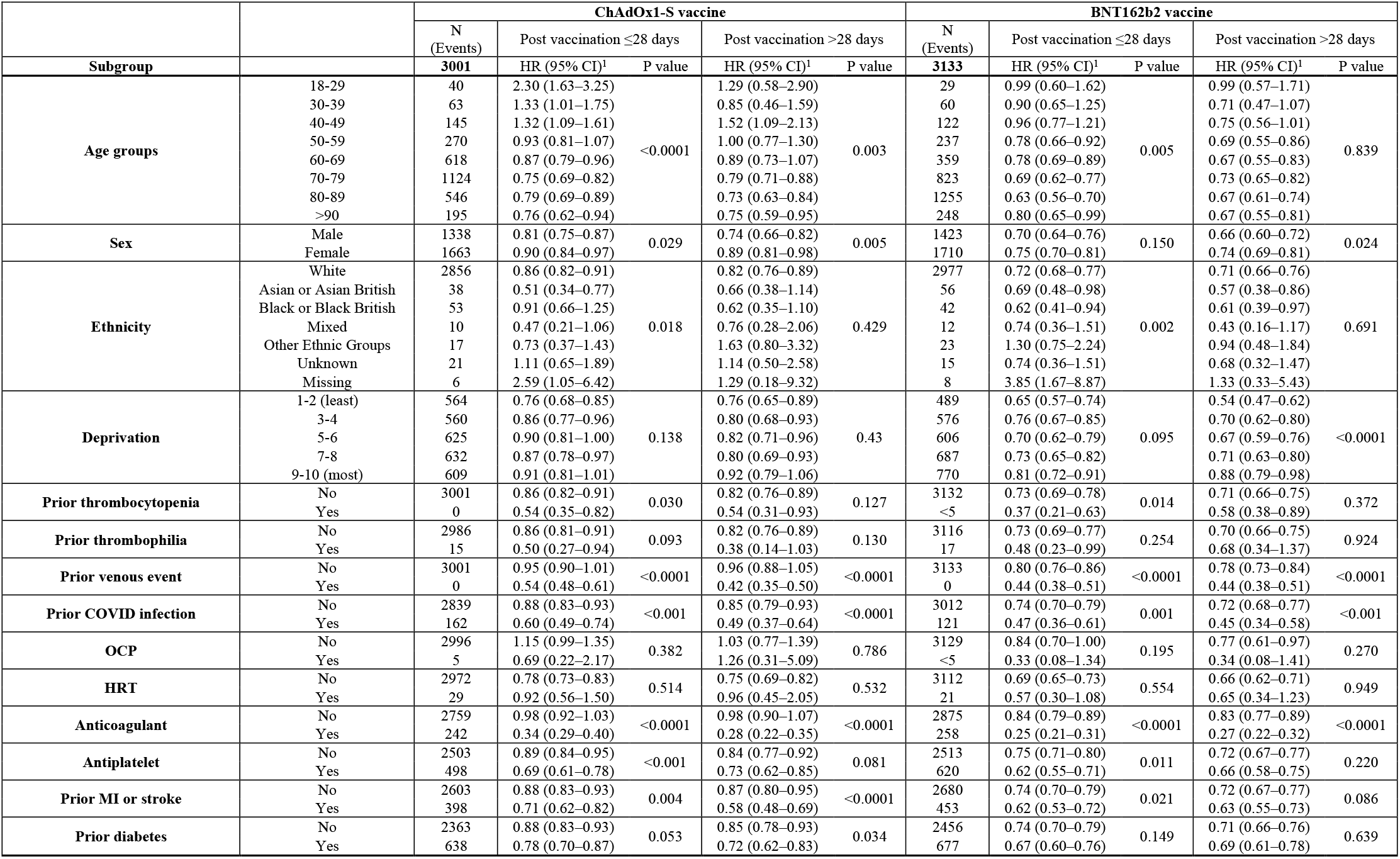
Fully adjusted hazard ratios (HRs) for the association of ChAdOx1-S and BNT162b2 with venous events 1-28 and >28 days after vaccination, within pre-specified subgroups. P values for between subgroup differences were derived using Wald tests. OCP: oral contraceptive. HRT: hormone replacement therapy.

**Supplementary table 2 (b).** Fully adjusted hazard ratios (HRs) for the association of ChAdOx1-S and BNT162b2 with arterial events 1-28 and >28 days after vaccination, within pre-specified subgroups. P values for between subgroup differences were derived using Wald tests. OCP: oral contraceptive. HRT: hormone replacement therapy.

|  |  | ChAdOx1-S vaccine |  |  |  | BNT162b2 vaccine |  |  |  |  |  |
| --- | --- | --- | --- | --- | --- | --- | --- | --- | --- | --- | --- |
|  |  | N<br>(Events) | Post vaccination ≤28 days |  | Post vaccination >28 days |  | N<br>(Events) | Post vaccination ≤28 days |  | Post vaccination >28 days |  |
| Subgroup | All arterial | 12890 | HR (95% CI)<sup>1</sup> | P value | HR (95% CI)<sup>1</sup> | P value | 16759 | HR (95% CI)<sup>1</sup> | P value | HR (95% CI)<sup>1</sup> | P value |
| Age groups | 18-29 | 12 | 1.72 (0.89–3.33) | <0.0001 | 1.84 (0.59–5.73) | 0.031 | 21 | 2.01 (1.13–3.57) | <0.0001 | 1.85 (0.96–3.59) | <0.0001 |
|  | 30-39 | 54 | 1.51 (1.11–2.06) |  | 1.54 (0.87–2.72) |  | 67 | 1.36 (1.00–1.86) |  | 1.09 (0.74–1.61) |  |
|  | 40-49 | 277 | 1.37 (1.20–1.57) |  | 1.29 (0.99–1.69) |  | 284 | 1.17 (1.01–1.36) |  | 0.86 (0.70–1.05) |  |
|  | 50-59 | 963 | 0.98 (0.91–1.05) |  | 0.89 (0.76–1.04) |  | 1062 | 1.06 (0.98–1.14) |  | 0.81 (0.72–0.91) |  |
|  | 60-69 | 2442 | 0.85 (0.81–0.90) |  | 0.90 (0.82–0.99) |  | 1705 | 0.89 (0.84–0.95) |  | 0.83 (0.76–0.92) |  |
|  | 70-79 | 5030 | 0.90 (0.86–0.94) |  | 0.93 (0.88–0.99) |  | 3993 | 0.85 (0.81–0.89) |  | 0.91 (0.86–0.96) |  |
|  | 80-89 | 2826 | 0.86 (0.81–0.91) |  | 0.88 (0.82–0.94) |  | 7733 | 0.74 (0.71–0.78) |  | 0.79 (0.76–0.82) |  |
|  | >90 | 1286 | 0.85 (0.78–0.93) |  | 0.86 (0.78–0.94) |  | 1894 | 0.72 (0.66–0.79) |  | 0.78 (0.73–0.84) |  |
| Sex | Male | 7116 | 0.78 (0.75–0.80) | <0.0001 | 0.75 (0.71–0.79) | <0.0001 | 9292 | 0.77 (0.74–0.79) | <0.0001 | 0.71 (0.68–0.73) | <0.0001 |
|  | Female | 5774 | 0.89 (0.86–0.93) |  | 0.87 (0.83–0.92) |  | 7467 | 0.85 (0.82–0.88) |  | 0.89 (0.85–0.92) |  |
| Ethnicity | White | 11729 | 0.82 (0.79–0.84) | <0.0001 | 0.80 (0.77–0.84) | 0.180 | 15492 | 0.80 (0.78–0.82) | <0.0001 | 0.79 (0.76–0.81) | <0.0001 |
|  | Asian or Asian British | 647 | 0.84 (0.76–0.93) |  | 0.74 (0.64–0.86) |  | 728 | 0.75 (0.67–0.84) |  | 0.63 (0.56–0.71) |  |
|  | Black or Black British | 217 | 0.90 (0.75–1.07) |  | 0.89 (0.70–1.15) |  | 190 | 0.77 (0.62–0.94) |  | 0.79 (0.64–0.99) |  |
|  | Mixed | 72 | 1.19 (0.90–1.57) |  | 0.56 (0.32–0.98) |  | 85 | 1.10 (0.82–1.49) |  | 0.81 (0.57–1.17) |  |
|  | Other Ethnic Groups | 113 | 0.93 (0.73–1.18) |  | 0.90 (0.64–1.28) |  | 121 | 0.89 (0.69–1.15) |  | 0.76 (0.57–1.01) |  |
|  | Unknown | 80 | 0.88 (0.67–1.16) |  | 0.75 (0.49–1.16) |  | 103 | 0.87 (0.64–1.18) |  | 1.09 (0.83–1.44) |  |
|  | Missing | 32 | 3.01 (1.98–4.57) |  | 1.91 (0.89–4.08) |  | 40 | 2.87 (1.81–4.56) |  | 2.87 (1.80–4.58) |  |
| Deprivation | 1-2 (least) | 2498 | 0.76 (0.71–0.80) | <0.001 | 0.71 (0.66–0.77) | <0.0001 | 2788 | 0.75 (0.71–0.79) | <0.0001 | 0.69 (0.65–0.73) | <0.0001 |
|  | 3-4 | 2514 | 0.81 (0.76–0.85) |  | 0.76 (0.71–0.82) |  | 2980 | 0.76 (0.72–0.81) |  | 0.74 (0.69–0.78) |  |
|  | 5-6 | 2683 | 0.85 (0.81–0.90) |  | 0.79 (0.74–0.85) |  | 3489 | 0.78 (0.74–0.83) |  | 0.79 (0.75–0.84) |  |
|  | 7-8 | 2616 | 0.83 (0.78–0.87) |  | 0.86 (0.80–0.92) |  | 3693 | 0.82 (0.78–0.87) |  | 0.83 (0.79–0.88) |  |
|  | 9-10 (most) | 2541 | 0.89 (0.85–0.95) |  | 0.88 (0.82–0.95) |  | 3772 | 0.90 (0.85–0.95) |  | 0.85 (0.80–0.89) |  |
| Prior thrombocytopenia | No | 12889 | 0.82 (0.80–0.85) | 0.334 | 0.80 (0.77–0.84) | 0.499 | 16758 | 0.80 (0.78–0.82) | 0.920 | 0.78 (0.76–0.81) | 0.136 |
|  | Yes | <5 | 0.75 (0.61–0.91) |  | 0.73 (0.57–0.95) |  | <5 | 0.81 (0.66–1.00) |  | 0.67 (0.54–0.82) |  |
| Prior thrombophilia | No | 12864 | 0.82 (0.80–0.85) | 0.059 | 0.80 (0.77–0.83) | 0.620 | 16735 | 0.80 (0.78–0.82) | 0.499 | 0.78 (0.76–0.81) | 0.098 |
|  | Yes | 26 | 0.48 (0.28–0.84) |  | 0.66 (0.32–1.39) |  | 24 | 0.66 (0.38–1.15) |  | 0.43 (0.21–0.88) |  |
| Prior venous event | No | 12874 | 0.83 (0.81–0.85) | 0.029 | 0.80 (0.77–0.84) | 0.504 | 16751 | 0.81 (0.78–0.83) | 0.022 | 0.79 (0.76–0.81) | 0.002 |
|  | Yes | 16 | 0.73 (0.66–0.82) |  | 0.77 (0.67–0.88) |  | 8 | 0.70 (0.63–0.79) |  | 0.66 (0.59–0.74) |  |
| Prior COVID infection | No | 12227 | 0.84 (0.81–0.86) | <0.0001 | 0.83 (0.79–0.86) | <0.0001 | 16234 | 0.81 (0.79–0.84) | <0.0001 | 0.80 (0.77–0.82) | <0.0001 |
|  | Yes | 663 | 0.63 (0.57–0.70) |  | 0.51 (0.45–0.59) |  | 525 | 0.57 (0.50–0.65) |  | 0.51 (0.45–0.59) |  |
| OCP | No | 12889 | 0.91 (0.79–1.05) | 0.983 | 0.89 (0.67–1.18) | <0.0001 | 16759 | 0.91 (0.79–1.06) | <0.0001 | 0.94 (0.75–1.16) | <0.0001 |
|  | Yes | <5 | 0.89 (0.12–6.51) |  | 0.00 (0.00–0.00) |  | 0 | 0.00 (0.00–0.00) |  | 0.00 (0.00–0.00) |  |
| HRT | No | 12832 | 0.85 (0.82–0.87) | 0.293 | 0.84 (0.80–0.88) | 0.820 | 16671 | 0.81 (0.78–0.83) | 0.061 | 0.79 (0.76–0.81) | 0.070 |
|  | Yes | 58 | 1.01 (0.73–1.40) |  | 0.79 (0.45–1.37) |  | 88 | 1.10 (0.79–1.51) |  | 1.08 (0.77–1.52) |  |
| Anticoagulant | No | 10726 | 0.84 (0.82–0.87) | <0.001 | 0.82 (0.78–0.85) | 0.015 | 13715 | 0.83 (0.81–0.86) | <0.0001 | 0.80 (0.77–0.83) | <0.001 |
|  | Yes | 2164 | 0.74 (0.69–0.79) |  | 0.74 (0.68–0.80) |  | 3044 | 0.66 (0.62–0.70) |  | 0.71 (0.67–0.76) |  |
| Antiplatelet | No | 7636 | 0.99 (0.96–1.03) | <0.0001 | 1.06 (1.01–1.11) | <0.0001 | 9770 | 0.96 (0.93–0.99) | <0.0001 | 1.05 (1.01–1.09) | <0.0001 |
|  | Yes | 5254 | 0.65 (0.63–0.68) |  | 0.59 (0.55–0.62) |  | 6989 | 0.65 (0.63–0.67) |  | 0.55 (0.53–0.57) |  |
| Prior MI or stroke | No | 6931 | 1.09 (1.05–1.13) | <0.0001 | 1.18 (1.12–1.24) | <0.0001 | 9428 | 1.08 (1.04–1.12) | <0.0001 | 1.22 (1.17–1.26) | <0.0001 |
|  | Yes | 5959 | 0.64 (0.61–0.66) |  | 0.60 (0.57–0.63) |  | 7331 | 0.60 (0.57–0.62) |  | 0.51 (0.49–0.53) |  |
| Prior diabetes | No | 8742 | 0.85 (0.83–0.88) | <0.0001 | 0.84 (0.80–0.88) | <0.001 | 11689 | 0.84 (0.82–0.87) | <0.0001 | 0.82 (0.79–0.85) | <0.0001 |
|  | Yes | 4148 | 0.77 (0.73–0.80) |  | 0.73 (0.69–0.78) |  | 5070 | 0.72 (0.69–0.76) |  | 0.70 (0.67–0.74) |  |

**Supplementary table 3.** Characteristics of patient who had an ICVT event before and after vaccination. Only percentage are presented, as disclosure control does not allow numbers <5 to be released.

|  |  | ICVT before vaccination % | ICVT after ChAdOx1, % | ICVT after BNT162b, % | p-value<sup>1</sup> |
| --- | --- | --- | --- | --- | --- |
| N |  | <210 | <40 | <20 | <0.0001 |
| Sex | Male | 40 | 33 | 21 | 0.24 |
|  | Female | 60 | 67 | 79 |  |
| Age | 18 - 29 | 19 | 17 | 0 | <0.0001 |
|  | 30 - 49 | 42 | 22 | 26 |  |
|  | 50 - 69 | 30 | 36 | 16 |  |
|  | 70 - 79 | 6 | 11 | 21 |  |
|  | 80+ | 3 | 14 | 37 |  |
| Ethnicity | Asian or Asian British | 15 | 6 | 0 | 0.38 |
|  | Black or Black British | 5 | 3 | 0 |  |
|  | Mixed | 2 | 0 | 0 |  |
|  | Other ethnic groups | 1 | 0 | 0 |  |
|  | White | 75 | 92 | 100 |  |
|  | Unknown or missing | 1 | 0 | 0 |  |
| Deprivation<sup>2</sup> | 1 - 2 | 29 | 17 | 11 | 0.19 |
|  | 3 - 4 | 20 | 14 | 26 |  |
|  | 5 - 6 | 25 | 25 | 21 |  |
|  | 7 - 8 | 13 | 25 | 16 |  |
|  | 9 - 10 | 13 | 19 | 26 |  |
| Smoking status | Current | 19 | 28 | 11 | 0.5 |
|  | Former | 21 | 17 | 32 |  |
|  | Never | 57 | 56 | 58 |  |
| Medical history | Stroke | 5 | 6 | 11 | <0.0001 |
|  | MI | 3 | 6 | 0 | 0.01 |
|  | DVT or PE | 8 | 6 | 5 | <0.0001 |
|  | Thrombophilia | 1 | 0 | 0 | 0.05 |
|  | Coronavirus infection<sup>3</sup> | 6 | 6 | 0 | <0.0001 |
|  | Diabetes | 10 | 17 | 37 | <0.0001 |
|  | Depression | 26 | 36 | 16 | <0.0001 |
|  | Obesity | 37 | 31 | 42 | <0.0001 |
|  | Cancer | 17 | 28 | 37 | <0.0001 |
|  | COPD | 2 | 11 | 16 | 0.78 |
|  | Liver disease | <1 | 3 | 0 | 0.61 |
|  | CKD | 5 | 11 | 32 | 0.25 |
|  | Major surgery<sup>4</sup> | 18 | 8 | 11 | <0.0001 |
|  | Dementia | 2 | 3 | 0 | 0.03 |
| Medications | Antiplatelet | 5 | 6 | 16 | 0.01 |
|  | BP lowering | 15 | 28 | 42 | <0.0001 |
|  | Lipid lowering | 12 | 22 | 26 | <0.0001 |
|  | Anticoagulant | 10 | 11 | 11 | <0.0001 |
|  | Oral contraceptive | 4 | 0 | 0 | <0.0001 |
|  | HRT | 1 | 0 | 0 | 0.05 |
| Number of diagnoses<sup>5</sup> | 0 | 71 | 50 | 58 | 0.04 |
|  | 1 - 5 | 29 | 50 | 42 |  |
| Number of medications | 0 | 31 | 11 | 2 | 0.02 |
|  | 1 - 5 | 62 | 83 | 5 |  |
|  | 6+ | 7 | 6 | 2 |  |

<sup>1</sup> Approximation if Chi squared test included one or more zero counts; <sup>2</sup> Index of Multiple Deprivation deciles where 1 indicates least deprived and 10 indicates most deprived; <sup>3</sup> After 31/12/2019 and prior to 08/12/2020; <sup>4</sup> In the last year; <sup>5</sup> The category '6+' contained no individuals and so was excluded.

**Supplementary figure 1:**
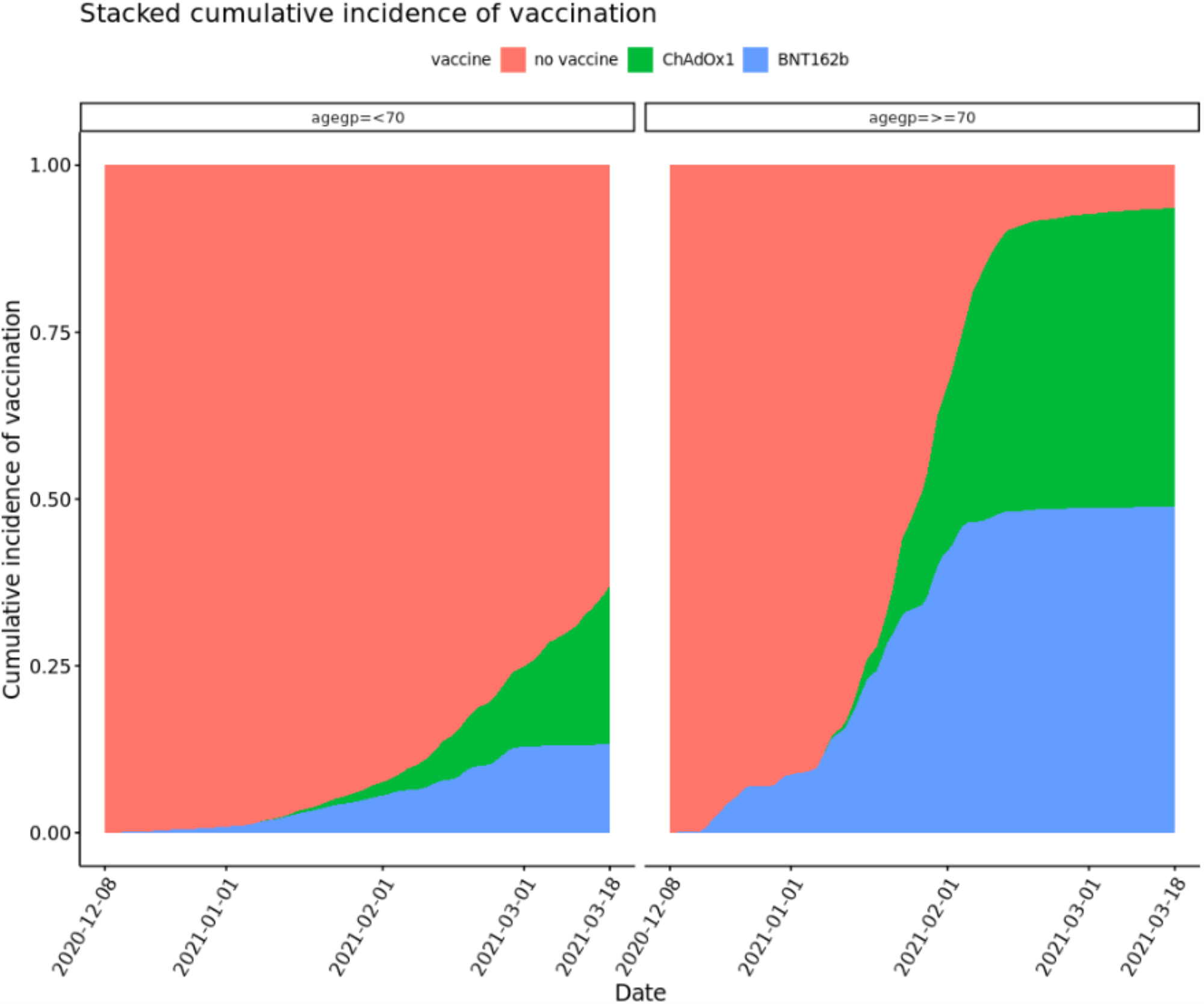
Cumulative incidence of vaccines of different types during follow up.

**Supplementary figure 2:**
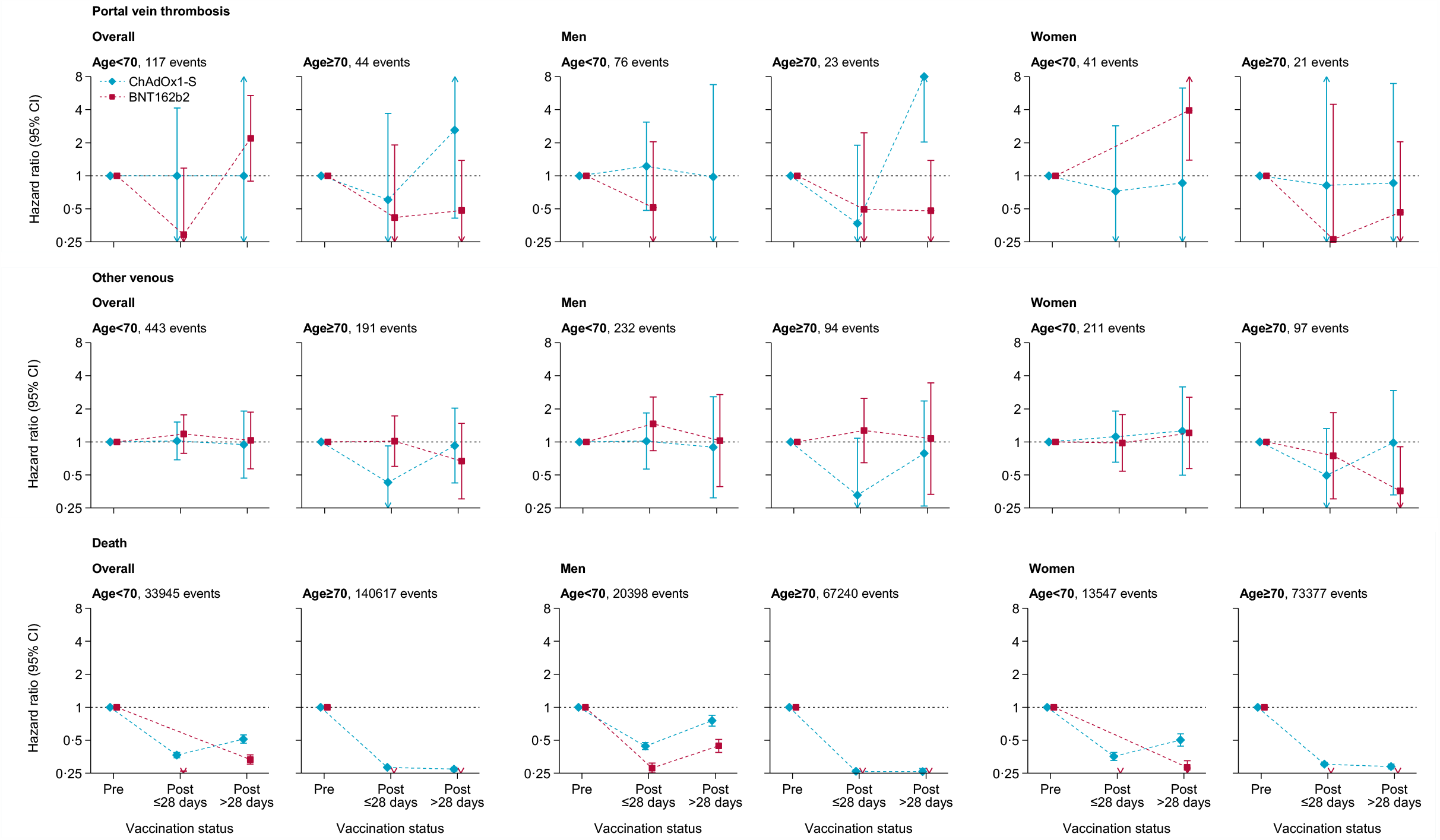
Adjusted hazard ratios for portal vein thrombosis, other venous events, and death after ChAdOx1-S or BNT162b2 vaccine.

## Notes

### Competing Interest Statement

The authors have declared no competing interest.

### Clinical Protocols

https://github.com/BHFDSC/CCU002_02

### Summary of Updates

Minor table changes

